# Potential for person-to-person transmission of henipaviruses: A systematic review of the literature

**DOI:** 10.1101/2023.02.26.23286473

**Authors:** Sonia Hegde, Kyu Han Lee, Ashley Styczynski, Forrest K Jones, Isabella Gomes, Pritimoy Das, Emily S. Gurley

**Affiliations:** Department of Epidemiology, Bloomberg School of Public Health, Johns Hopkins University, Baltimore, MD, USA; Division of Infectious Diseases & Geographic Medicine, Stanford University, Stanford, CA, USA; Institute of Health and Wellbeing, Federation University Australia, Ballarat, Victoria, Australia

## Abstract

Nipah virus – Bangladesh (NiV_B_) is a bat-borne zoonosis transmitted between people through the respiratory route, posing a pandemic risk. The risk posed by related henipaviruses, including Hendra virus (HeV) and Nipah virus – Malaysia (NiV_M_) is less clear. We conducted a broad search of the literature encompassing both human infections and animal models to synthesize evidence about potential for person-to-person spread of these henipaviruses. More than 600 human infections have been reported in the literature, but information about biological processes related to transmission is limited; information on viral shedding was only available for 40 case-patients. There is substantial evidence demonstrating person-to-person transmission of NiV_B_, though there is also evidence that NiV_M_ has been transmitted from person to person. Less direct evidence is available about the risk for person-to-person transmission of HeV, but animals infected with HeV shed more virus in the respiratory tract than those infected with NiV_M_ suggesting potential for transmission. As the family of known henipaviruses continues to grow, shared protocols for conducting and reporting from human investigations and animal experiments are urgently needed to advance our understanding of transmission risk.

## Introduction

Zoonotic pathogens with demonstrated ability to infect and transmit between people are potential pandemic threats. In 2015, the World Health Organization named Nipah virus as one of the most dangerous emerging zoonotic disease threats because of its high case fatality and ability to transmit from person-to-person^1^. Nipah virus belongs to the genus *Henipavirus*, in the family *Paramyxoviridae*, along with Hendra virus, which has caused spillovers into horses in Australia and has also caused human infections with severe clinical outcomes^2, 3^. Other henipaviruses, including Cedar virus, Mojiang virus, and Langya virus, have also been identified, but are much less common. The natural reservoirs for henipaviruses are pteropid bats, which are large fruit bats whose habitats span from South and Southeast Asia to East Africa and Australia.

Nipah virus outbreaks in Bangladesh and India have been reported since 2001 and have regularly been associated with person-to-person transmission^1, 2^. Evidence suggests that transmission occurs through close contact with patients and their respiratory secretions^1^. However, person-to-person transmission was noted in the first outbreak identified in Malaysia and Singapore in 1998 and 1999^2, 4^, where spillover into pigs and transmission to abattoir workers were salient features. While very similar, the Nipah virus strain identified in Bangladesh and India differed phylogenetically from the virus strain identified in the Malaysia and Singapore outbreak^5^. Hendra virus has not been associated with person-to-person transmission, and is distinct from Nipah virus; partial N gene fragments can be used to genotype viruses at the major clade level (i.e. Hendra vs. Nipah Malaysia vs. Nipah India/Bangladesh), but cannot mimic full-genome genetic variation^5^. Epidemiologic evidence about person-to-person transmission across countries has led to the suggestion that differences in transmissibility might be driven by genetic differences in henipavirus strains^6^ and that the India/Bangladesh Nipah strain is better adapted to human spread than other henipaviruses. However, an outbreak of henipavirus in the Philippines in 2015 was associated with person-to-person transmission and the virus implicated in the outbreak was more closely related to the Malaysian Nipah strain than the South Asian strain^7^.

Understanding which henipaviruses have the greatest potential for person-to-person spread would improve our scientific understanding of their pandemic risk. If the India/Bangladesh Nipah strain poses the greatest risk for pandemic spread among known henipavirus strains, then resources should be targeted specifically to that geographic region to reduce pandemic risk. If the differences among known strains are merely an artifact of our epidemiologic observations rather than true variation in risk, then prevention strategies must address this broader henipavirus risk. We conducted a systematic review and meta analysis of human epidemiologic and clinical studies and studies of infections in mammals to compare transmission potential among the three henipaviruses known to infect humans: the India/Bangladesh strain of Nipah (NiV_B_), the Malaysian strain of Nipah (NiV_M_), and Hendra virus (HeV).

## Methods

### Literature search and study selection

We conducted a broad search of the literature using PubMed, Embase, Cochrane Central Register of Controlled Trials, Web of Science, and Scopus. Gray literature was searched using IndMED, KoreaMED, and WHO Global Index Medicus. Search strategies (Supplementary Table 1) were used to identify publications indexed by May 30, 2019 using the genus (“Henipavirus”) and species names (“Hendra” and “Nipah”). We reviewed the references of selected papers and identified 2 studies published prior to the use of ‘Hendra’, when it was referred to as ‘Equine morbillivirus’, that we included in the review, and 1 study on non-human primates which was published on May 29, 2019 but was not yet indexed.

Titles and abstracts were independently reviewed by two members of the study team and excluded if they lacked primary data, were an *in vitro* experiment, or were in languages other than English, French, Spanish, and Dutch. We also excluded studies of infection in bats as these do not provide evidence of potential human-to-human spread. Studies were included for data extraction if two team members reviewed the full text and confirmed that the study included human studies with primary data on secondary attack rates or transmission between people, or viral shedding (from any biological specimen) or viremia in either humans or experimentally infected mammals. Any discrepancies between reviewers were resolved through discussion between all reviewers.

### Data extraction and analysis

For human studies, we extracted case-patient (or confirmed case) data including demographics, incubation period, symptomatology, whether or not the case was infected through person-to-person transmission, and clinical outcome. When available, we also extracted data on viral shedding by day post-illness onset.

We calculated the proportion of case-patients that experienced respiratory symptoms, median incubation period, case-fatality, and secondary attack rates and compared these metrics between henipaviruses. We defined respiratory symptoms as those case-patients that either were explicitly described to have respiratory distress, difficulty breathing, or “influenza-like illness (ILI) and encephalitis”, as some articles described case-patients with acute encephalitic syndrome as experiencing respiratory distress. We described the number of secondary cases for all Nipah virus cases where this information was published. We graphed the proportion of patients who shed virus in respiratory secretions by days since symptom onset using available data.

Information about viral shedding in humans was very rare so we relied primarily on animal studies to characterize and compare viral shedding. For experimental animal studies, we extracted data on animal species, route and dose of inoculation, and viral shedding by type of biological specimen collected and day post-inoculation. For observational studies of naturally infected animals, we extracted any information about viral shedding and type of biological specimen tested.

We identified the studies that used the same animal model, dose, and comparable route of inoculation but with different henipavirus strains to allow us to make comparisons of key characteristics of transmission potential, including quantity and duration of viral shedding. As exposure to respiratory secretions is considered the primary route of person-to-person transmission^1^, we focused our analysis on animal viral shedding from the respiratory tract. We examined the virus quantity (in genome equivalents) shed post inoculation in oral and nasal samples as a proxy measure of infectiousness for person-to-person transmission^8, 9^, and we examined the infectious period, as determined by polymerase chain reaction (PCR) or real-time polymerase chain reaction (RT-PCR) viral detection in the days post inoculation, as a proxy measure of the duration of viral shedding. We also examined the duration of viral shedding prior to the onset of respiratory signs and the peak viral quantity both before and during respiratory signs.

## Results

We identified 2,465 studies that met our inclusion criteria for review (Supplementary Figure 1). After review, 52 human and 78 animal studies met the inclusion criteria for data extraction. The number of published studies increased over time, primarily among animal studies (Supplementary Figure 2).

### Human studies

We identified 52 studies with data on confirmed human henipavirus infections; 6 on Hendra virus in Australia and the remaining 46 on Nipah virus: 14 from Malaysia, 3 from Singapore, 1 from the Philippines (identified as Nipah-like virus), 19 from Bangladesh, and 9 from India (Supplementary Table 2). Twenty-nine studies (56%) investigated person-to-person transmission and 19 (37%) investigated viral shedding in oral, urine, or semen samples. Eleven studies (21%) investigated both transmission and viral shedding, and all such studies were from Bangladesh, India, and the Philippines.

#### Transmission potential of HeV

Only 6 human cases of HeV infection have been reported in the published literature^10–12^, though 7 total human cases of HeV infection have occurred since 1994^13^, and none of these studies explicitly investigated person-to-person transmission. The presumptive exposure route was through contact with a sick or dead horse infected with HeV. However, one case of HeV had exposure to both an infected horse and a confirmed human case approximately 4 days prior to illness onset such that person-to-person transmission could not be ruled out^14^. The case in question developed symptoms 11 days after exposure to an infected horse (performed a nasal cavity lavage during the last 3 days of the horse’s incubation period) and was exposed in the work-place to the index case during his incubation period and first two days of illness. The index case developed symptoms 9 days after exposure to the same infected horse during the same procedure and presented with symptoms to the clinic 4 days prior to the case in question^14^. Among the 6 cases, 50% had respiratory symptoms and 50% died (Table 2); the seventh HeV case also died^13^. Three HeV cases from the 2004 and 2008 HeV outbreaks had oral or nasal and urine specimens collected to look for viral shedding (a total of 10 samples among the 3 case-patients were tested). Two of the 3 cases had evidence of HeV RNA by PCR within 15 days post illness onset (1 patient from oral/nasal specimen and 1 patient from urine specimen) both of whom also had signs of respiratory distress; the third case only had 1 sample taken 365 days post illness onset, which was negative. One case-patient had urine samples at days 23 and 30 post illness onset that had evidence of HeV RNA by PCR. There was no investigation of asymptomatic infections among exposed humans, though there was an investigation and evidence of asymptomatic infection among exposed horses^11, 15^.

#### Transmission potential of NiV_M_

Studies on the transmission potential of NiV_M_ were conducted in Malaysia, Singapore, and the Philippines, representing a total of 294 symptomatic human cases and 308 total infections (Table 2). Of the 18 studies on NiV_M_ in humans, 4 investigated person-to-person transmission. In Malaysia and Singapore, although no healthcare workers who cared for hospitalized outbreak-related patients or abattoir workers (as the outbreak occurred near pig farms) developed symptoms suggestive of NiV_M_ infection, they were investigated to identify evidence of IgG antibodies against NiV_M_^4, 16^. Three nurses in Malaysia and 22 healthcare workers in Singapore^16^ had IgG antibody responses, though none of them had neutralizing antibodies. Ten abattoir workers (0.7%; 10/1469) were identified as having asymptomatic infections, however, the exposure was assumed to be infected pigs rather than infected humans^16^. Investigators of these outbreaks did not investigate person-to-person transmission to family caregivers, and the paper from Singapore specifically stated that transmission within households was not investigated because of concerns of inciting panic^16^. Further, as the outbreak was primarily investigated retrospectively, investigations of person-to-person transmission within households or family contacts of patients would have been very logistically difficult to do. The only evidence reported of transmission in households during the outbreak in Malaysia came from a report of an episode of late-onset encephalitis in a woman who did not live in the outbreak area, but who had traveled to the area during the outbreak to care for her aunt who was a Nipah case-patient. The woman was diagnosed with late-onset Nipah encephalitis 11 years after the initial outbreak^17^.

In the outbreak in the Philippines, 17 cases were identified. Seven cases slaughtered horses or consumed horse meat and 5 cases (29%) were exposed to other human cases but not to any horses. At least 12% (≥2/17) of case-patients infected one other person; based on history of patient contacts, 5 secondary cases were caused by person-to-person transmission from a minimum of 2 cases (Table 1; Figure 1).

**Figure 1.**
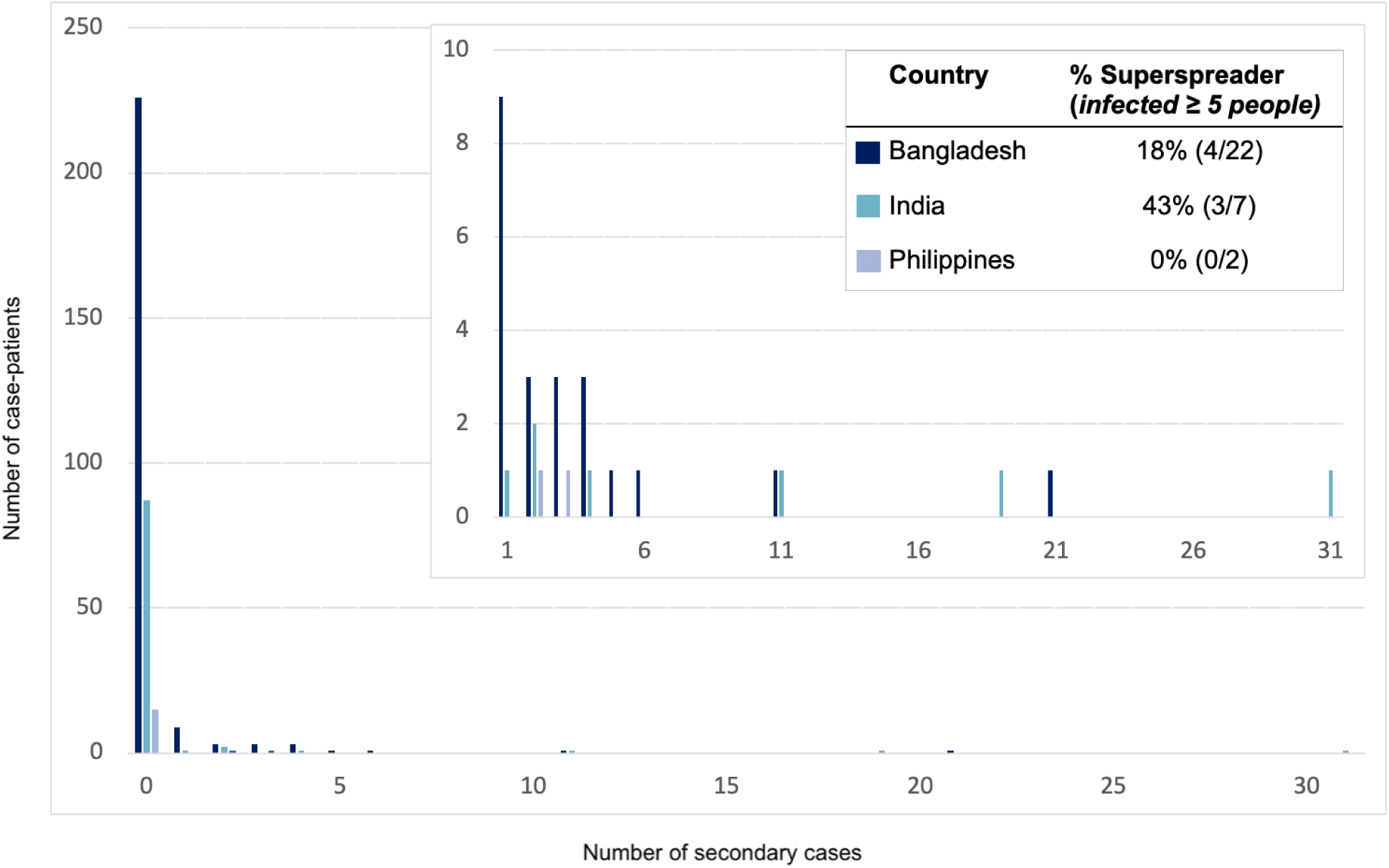
The distribution of secondary cases per Nipah case-patient (offspring distribution) and the proportion of those who transmitted who were superspreaders in countries where any person-to-person transmission of henipaviruses has been observed – Bangladesh, India, and the Philippines, 2001 - 2018.

**Table 1.**
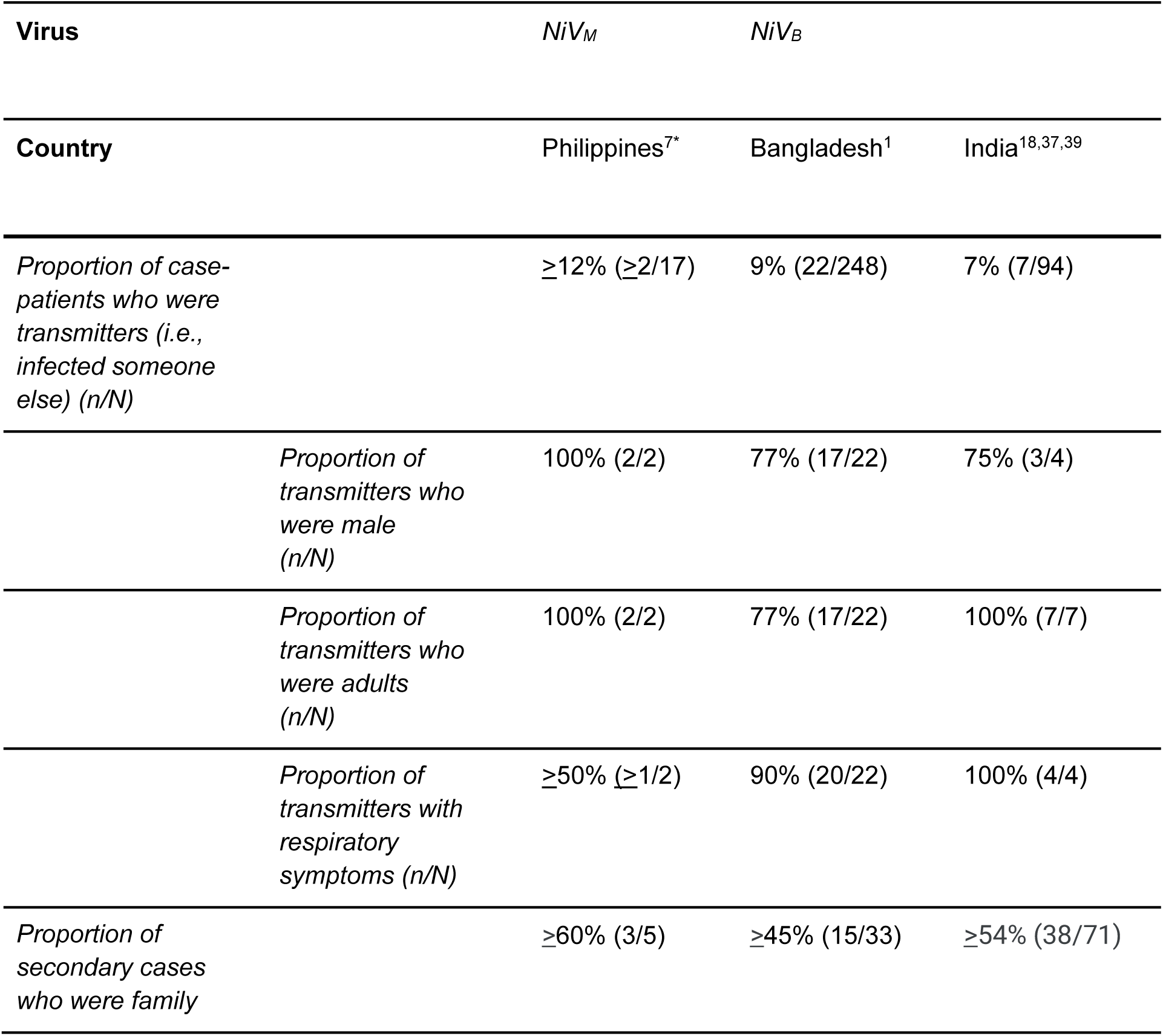
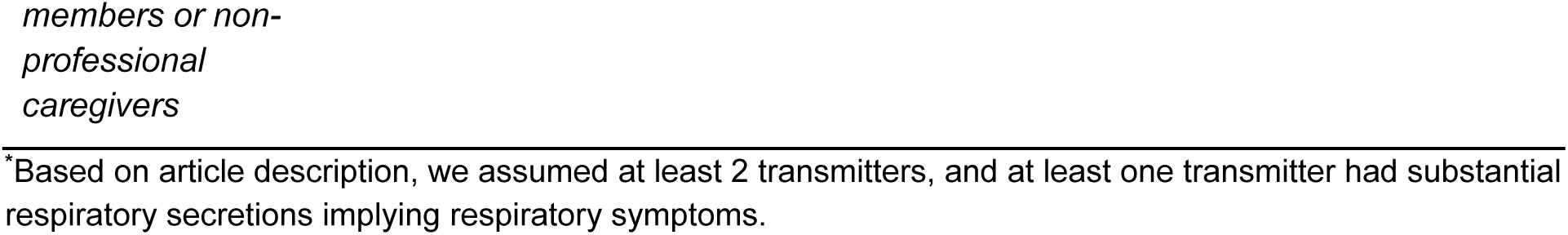
Description of person-to-person henipavirus transmission and characteristics of case-patients who infected others by henipavirus strain and country in the subset of studies where person-to-person transmission was identified, 2001 - 2018. All estimates below are based on the studies where relevant information was provided.

No patients infected with NiV_M_ had any samples collected to investigate viral shedding. While 23% (32/138) of patients identified in the outbreaks in Malaysia and Singapore had respiratory symptoms, 94% (16/17) of the patients in the Philippines outbreak did (Table 2).

**Table 2.**
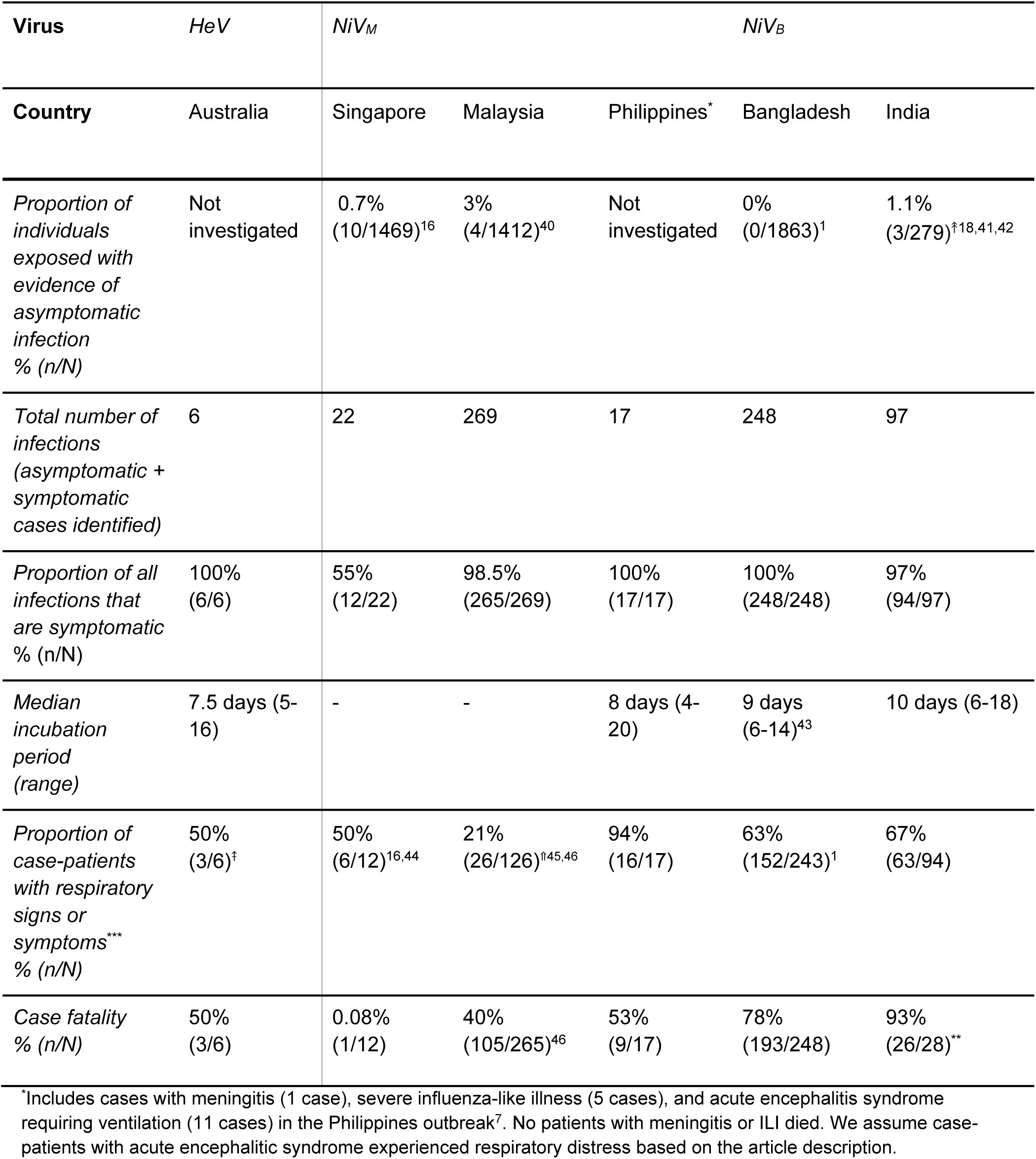
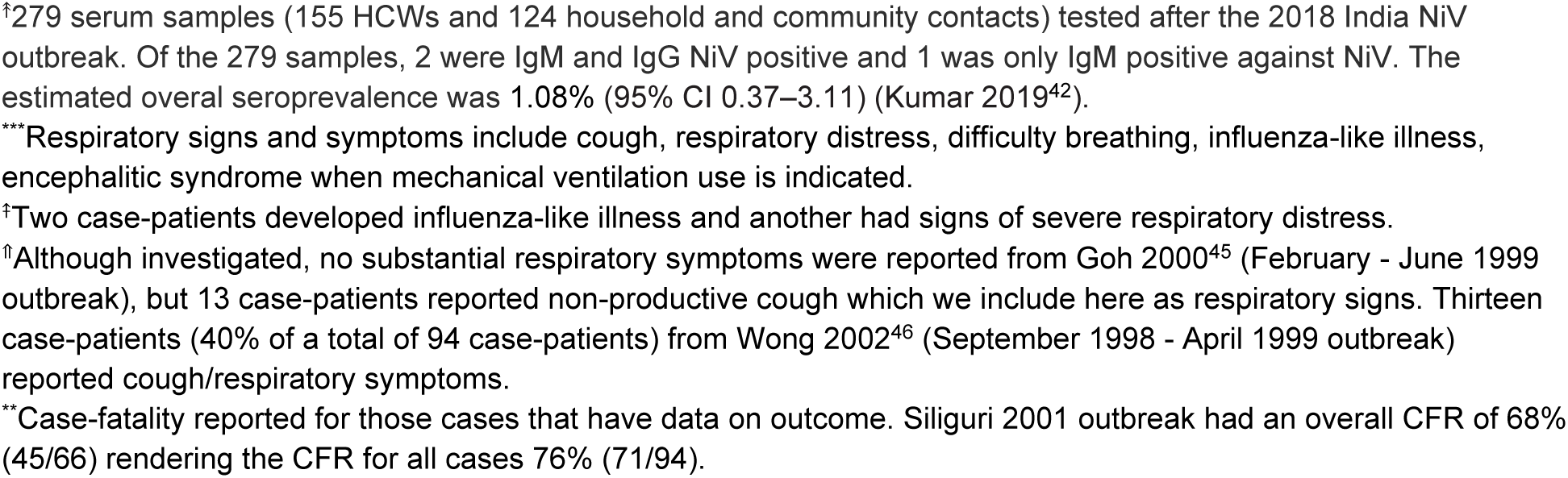
The natural history of infection for henipaviruses (Hendra virus: HeV, Nipah virus - Malaysia: NiV_M_, and Nipah virus - India/Bangladesh: NiV_B_) by country of outbreak origin using data from outbreaks identified during the 2001-2018 time period. References are cited to indicate from where data was used to derive estimates for each metric.

#### Transmission potential of NiV_B_

All 28 studies of NiV_B_ outbreaks in Bangladesh and India, representing 342 total case-patients and 345 total infections (3 asymptomatic infections were found during a serological survey among patient contacts after the 2018 outbreak in India), have investigated the possibility of person-to-person transmission and secondary cases have been regularly identified. Twenty-nine case patients infected at least 1 other person (transmitter); 7% (7/94) of case-patients from India and 9% (22/248) of case patients from Bangladesh (Table 1). Among the 29 NiV_B_ transmitters in the published literature, all were adults and 77% (20/26 where data was available) were males. Ninety-two percent of transmitters (24/26 with available data) had respiratory symptoms and all died.

Among case-patients who transmitted NiV_B_, 34% (10/29) transmitted to just 1 person and 24% (7/29) infected ≥5 other people; the size of outbreaks was heavily influenced by a few individuals who transmitted the virus to others as 8% of case-patients were responsible for the majority of transmission events (Figure 1). Roughly half (51%) of all 149 secondary cases were family members or non-professional caregivers and 9% were healthcare workers. In the 2018 India outbreak, during which detailed data was recorded, 22 of 23 total cases were a result of nosocomial transmission in 3 different hospitals.^18^ The longest transmission chain reported was from a 2004 outbreak in Bangladesh, where 5 generations of transmission occurred^19^.

Seventeen patients from Bangladesh and 20 from India were investigated to identify viral shedding; 37 unique case-patients from the 2008 and 2013-2014 Bangladesh and 2001, 2007, and 2018 India Nipah virus outbreaks provided 55 unique samples including urine, oral/nasal, and semen samples. One study from the 2018 Nipah virus outbreak in India provided PCR cycle threshold (Ct) values to quantify viral load^20^, and one other study from Bangladesh^1^ provided Ct values from throat swabs from case-patients, though no epidemiologic data accompanied these viral load data. There are 27 patients who contributed at least 1 oral or nasal swab sample in the published literature (total 37 samples), and 25 patients (total 32 samples) contributed sampled during their first week of illness. Seventy-six percent (28/37) of respiratory samples that were tested had evidence of viral RNA, and 81% (26/32) had evidence of viral RNA in the first week of illness. Respiratory samples were collected an average of 7.6 days post illness onset and samples that were positive were collected an average of 5.2 days after illness onset. More respiratory samples were collected 7 days post onset than other days (n=8) and all but 1 had evidence of NiV RNA by PCR (Figure 2). Only 1 patient was tested for viral RNA in semen and was found to be positive on days 16 and 26 but negative on days 42 and 59.

**Figure 2.**
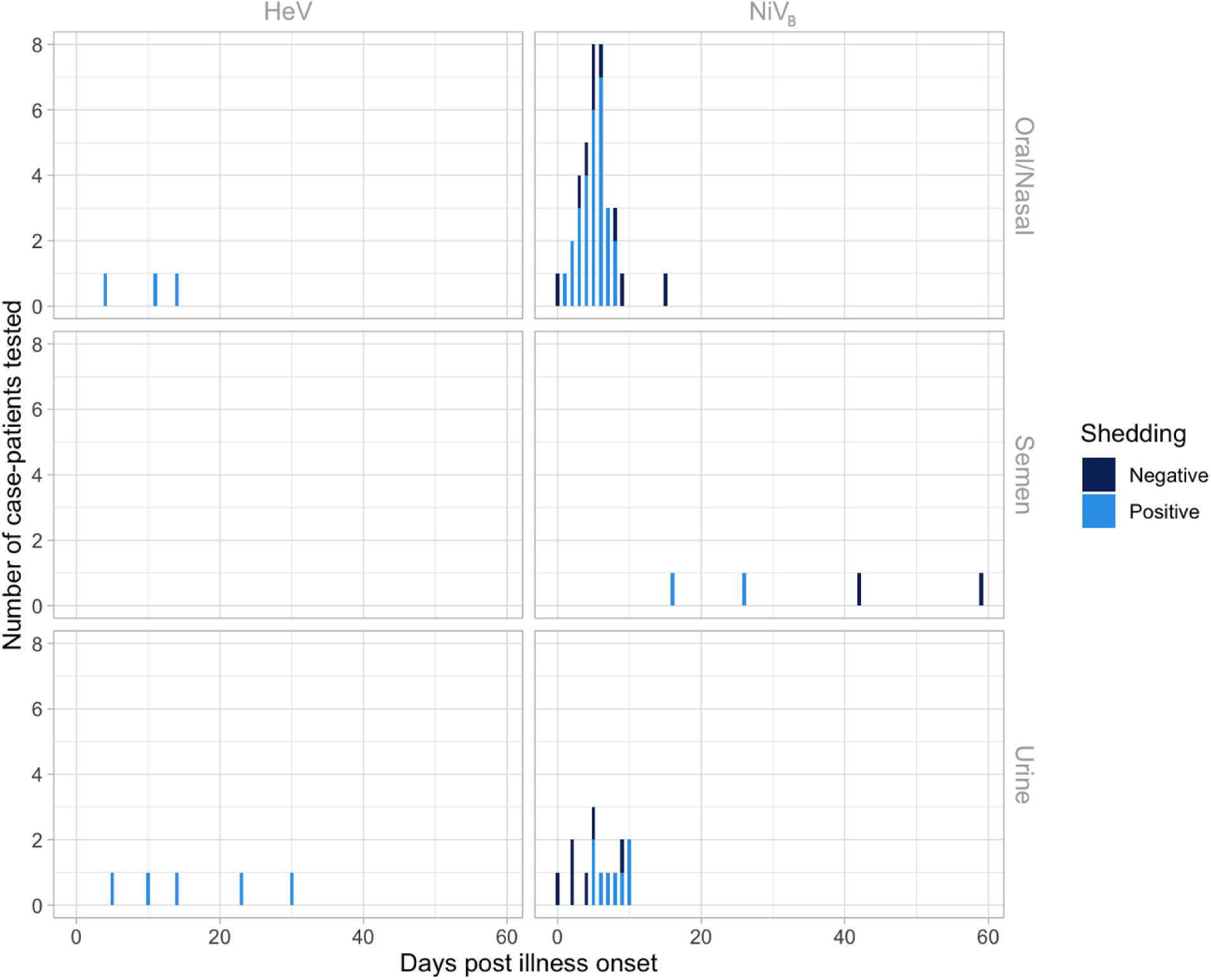
Evidence for henipavirus viral shedding in (a) 8 human oral/nasal and urine samples by days post illness onset using polymerase chain reaction (PCR) testing for 3 unique case-patients with confirmed Hendra virus (HeV) infection from the 2004 and 2008 HeV outbreaks in Australia, and (b) 55 human oral/nasal, semen and urine samples by days post illness onset using PCR testing for 37 unique case-patients with confirmed Nipah virus (NiV_B_) infection from the 2008 and 2013-2014 Bangladesh and 2001, 2007, and 2018 India NiV outbreaks. Though not shown in this figure, two additional urine samples from case-patients with HeV were PCR tested at days 365 (1 case-patient from the 2004 HeV outbreak) and 548 (1 case-patient from the 2008 HeV outbreak) post illness onset. Both of these urine samples were PCR negative.

In Bangladesh, 1 study summarizing case findings from 14 years of data found contacts of Nipah patients were significantly more likely to be infected if they had a longer duration of exposure to the patient (>12 hours), or if they had contact with the patient’s body fluids, particularly respiratory secretions^1^. Contacts also reported close contact with Nipah case-patients towards the end of life in Bangladesh^21^. Anecdotal evidence from the Kerala, India outbreak in 2018 suggests a similar pattern where contact nearer to the day of death resulted in transmission more often than contact earlier in the course of illness^18^.

For Nipah case-patients that died in Bangladesh and India and that we have information on time of symptom onset and death (N=37), the median number of days from symptom onset to death was 6 days (IQR 5-7.5) for those that transmitted the virus to another person (mean 6.7 (3.5-13) days; N=24) compared to 7 (IQR 6 - 9.5) days for case-patients that died but did not transmit (mean 8.9 (3-31) days; N=39).

### Animal studies

We identified 78 animal studies with primary data on the amount or duration of viral shedding, encompassing 11 animal model types: non-human primate, ferret, hamster, horse, pig, mouse, cat, dog, guinea pig, rat, and shrew (Supplementary Table 3). Thirty-six studies investigated infections with NiV_M_, 21 with HeV, 6 with NiV_B_, 1 with Cedar virus^22^ and 1 with Mojiang virus^23^. Six studies directly compared animals infected with NiV_M_ and NiV_B_, five compared NiV_M_ and HeV, and one compared NiV_M_, NiV_B_ and HeV. The Nipah virus strains used in animal studies were derived from a case-patient from the 2004 Rajbari, Bangladesh outbreak^19^ (NiV_B_) and a case patient from 1999 Malaysia outbreak among pig farmers^24^ (NiV_M_). The Hendra virus strains used in animal studies were derived from infected horses from the 1994^15^ and 2008^25^ outbreaks in Brisbane, Australia.

The route and dose of inoculation and types of biological samples collected varied substantially between study protocols, even within the same animal model, which made inferences about differences between viruses difficult (Supplementary Table 3). Given the heterogeneity in study protocols and considering the primary route of person-to-person transmission is exposure to respiratory secretions^1^, we analyzed data from studies with PCR or RT-PCR results from oral, nasal, and/or nasopharyngeal samples that directly compared 2 or more henipavirus strains within the same experiment or that only inoculated animals with 1 strain but used nearly identical methods with at least 1 other study (Supplementary Figure 3). Ultimately, we limited our analysis to 8 studies where the same methods were used for more than 1 virus within the same animal model to allow for equivalent comparisons: 4 African green monkey and 4 ferret studies.

#### African green monkey studies

The 4 African green monkey studies^26–29^ included in the analysis were conducted by the same research group and used nearly identical protocols, though the route of inoculation differed slightly across studies (Mire et al 2014^26^ and Geisbert et al^27^ inoculated intratracheally, Mire et al 2016^28^ and Mire et al 2019^29^ inoculated intranasally and intratracheally). In all, 20 animals were inoculated with 10^5^ plaque-forming units (PFU) with 3 different henipavirus strains: 7 with NiV_B_, 9 with NiV_M_, and 4 with HeV.

All animals inoculated with HeV and more than half of the animals inoculated with NiV_M_ were first investigated for viral shedding 3 days post inoculation (Figure 4). One of 4 animals (25%) inoculated with HeV began shedding on day 3 post-inoculation and 50% were shedding by day 5; 67% (6/9) of animals inoculated with NiV_M_ and 57% (4/7) of animals inoculated with NiV_B_ were shedding by day 5. Shedding often occurred prior to the onset of respiratory symptoms for animals with detectable virus (59%, 10/17) (Figure 4, Table 3). Viral shedding typically continued until the animal reached criteria for euthanasia (Figure 4, Table 3). Time to euthanasia was longer for African green monkeys inoculated with NiV_M_ (mean 9.3 days, range 8-10 days) compared to NiV_B_ (7.3 days, 7-8 days) and HeV (8.0 days, 8-8 days), contributing to longer shedding durations in animals inoculated with NiV_M_ (4.8 days, 0-8 days) compared to NiV_B_ (2.8 days, 0-5 days) and HeV (3.0 days, 0-6 days).

**Figure 4.**
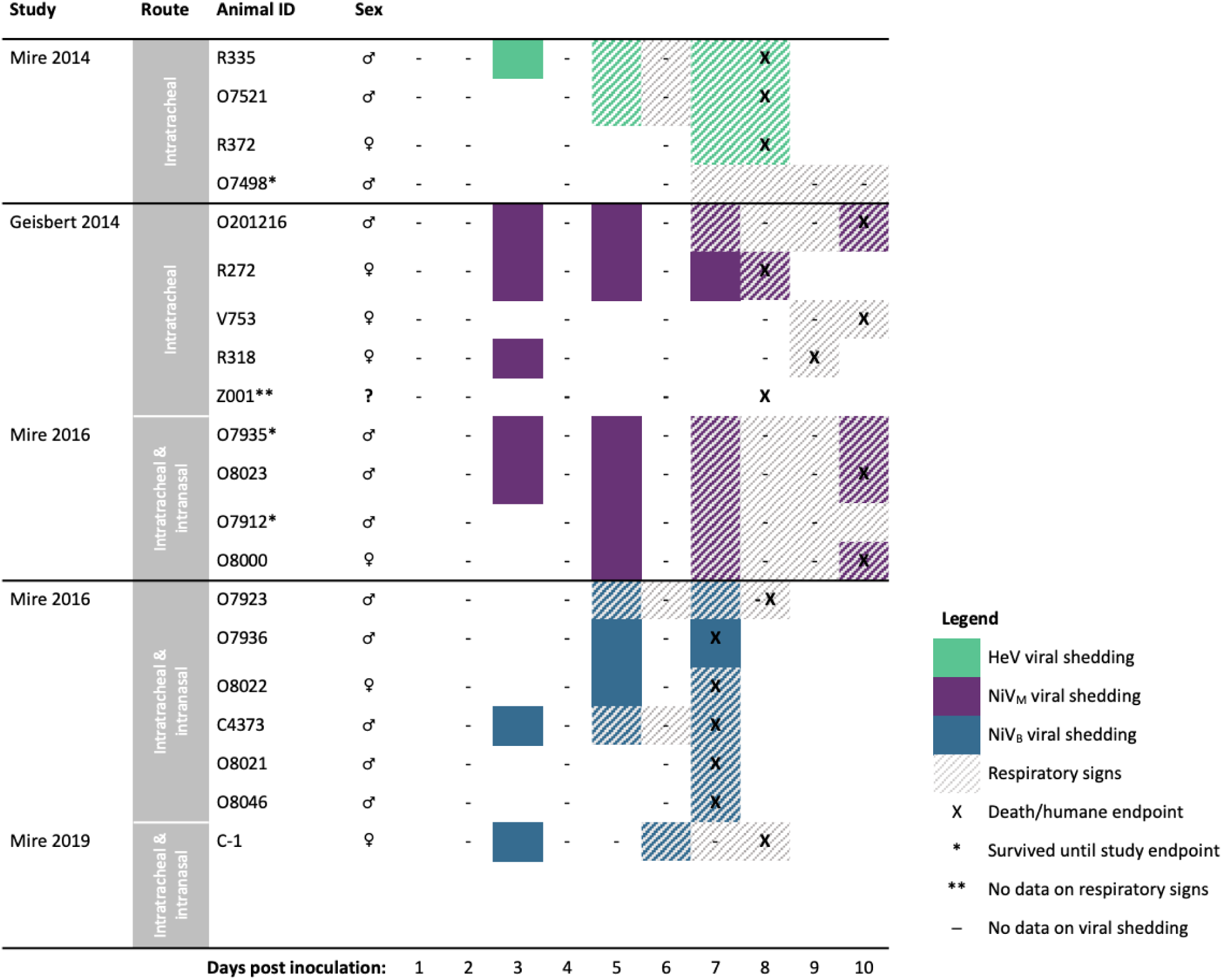
Comparing onset and duration of oral viral shedding and respiratory signs, and timing of humane endpoints among African green monkeys (N=20) infected with Hendra virus, Nipah virus Bangladesh (NiV_B_), or Nipah virus Malaysia (NiV_M_), by study. The studies used here, including the route of inoculation employed, are as follows: Mire 2014^26^ (intratracheal; N=4), Geisbert 2014^27^ (intratracheal; N=5), Mire 2016^28^ (intratracheal & intranasal; N=10), Mire 2019^29^ (intratracheal & intranasal; N=1).

**Table 3.**
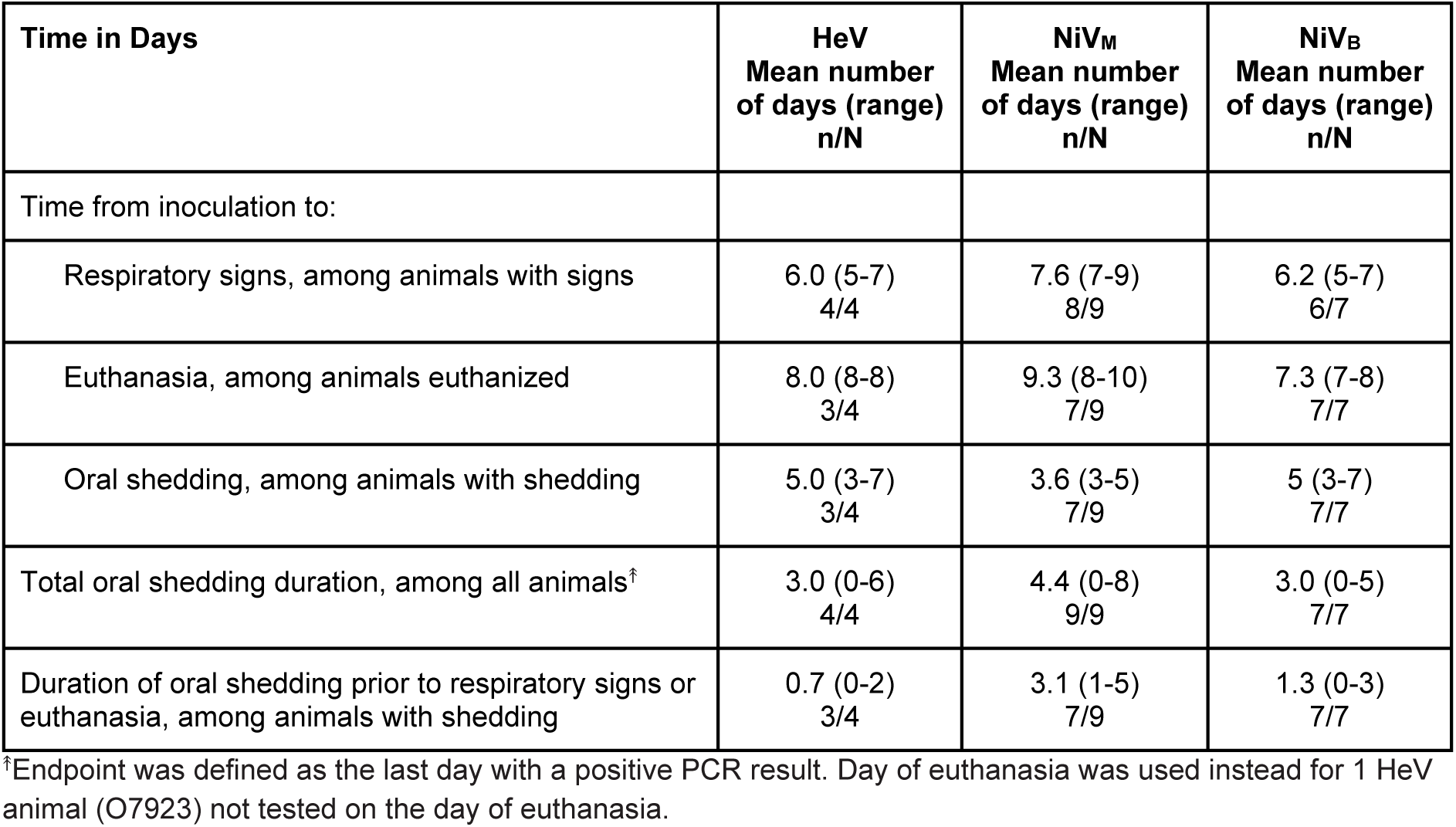
Comparing the trajectory of infection, including timing of viral shedding, respiratory signs, and euthanasia among African green monkeys infected with Hendra virus (HeV), Nipah virus Malaysia (NiV_M_), or Nipah virus Bangladesh (NiV_B_) in studies that used similar protocols. (N=20) The sample size (n/N) indicates how many animals provided data for the estimation of reported time in days.

African green monkeys inoculated with NiV_M_ developed respiratory signs later (mean 7.6 days, range 7-9) than those inoculated with NiV_B_ (6.2, 5-7) and HeV (6.0, 5-7) (Figure 4, Table 3). This resulted in longer periods of viral shedding prior to the onset of respiratory signs or euthanasia in the case of animals without signs. Animals inoculated with NiV_M_ shed virus for an average of 3.1 days (range 1-5) prior to signs, compared to 1.3 days (0-3) with NiV_B_ and 0.7 days (0-2) with HeV.

In oral samples, we observed a mean virus quantity over 100-fold higher starting on day 5 post inoculation in African green monkeys inoculated with NiV_B_ compared to NiV_M_ and HeV (Figure 5A). The same pattern was not observed in nasal samples, where we noted a similar rise in virus quantity for both NiV_B_ and NiV_M_ by day 1 post inoculation (Supplementary Figure 4). Among animals with HeV, we observed consistently lower virus quantities in nasal samples with 2 of 4 animals having no detectable virus up to 7 days post inoculation; however, all 4 animals inoculated with HeV were inoculated intratracheally, as opposed to both intratracheal and intranasal routes in other studies. For each virus strain, the mean virus quantity increased in oral samples after onset of respiratory symptoms (Figure 5B, 5D). Animals inoculated with NiV_B_ expressed the highest virus quantity in oral samples both before and during respiratory signs. There were no studies that reported on detection of culturable virus from oral or nasal specimens.

**Figure 5.**
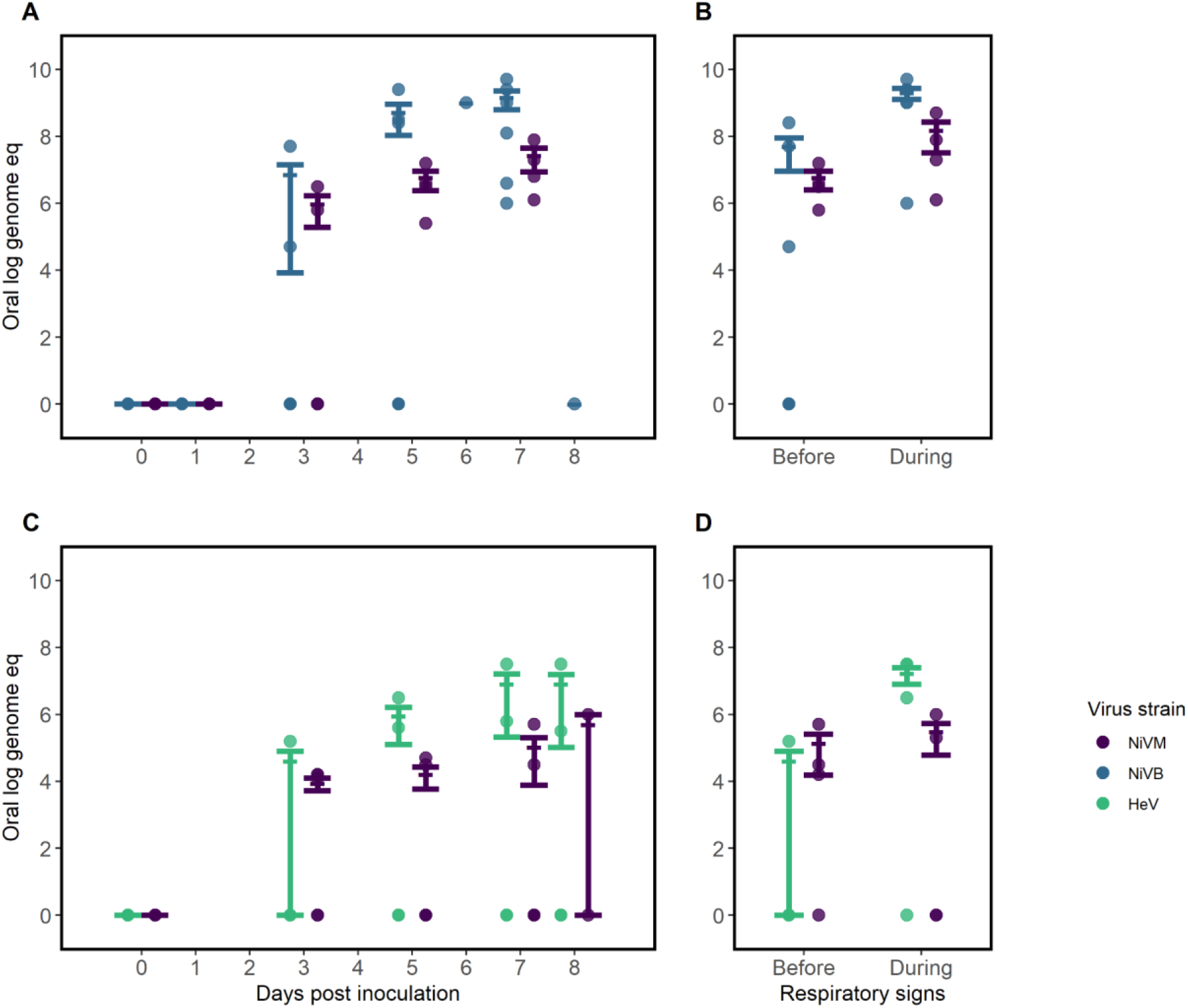
Comparing virus quantity in oral samples of African green monkeys infected with Hendra virus (HeV) or Nipah virus Bangladesh (NiV_B_) compared with Nipah virus Malaysia (NiV_M_) in studies with similar study protocols, by route of inoculation. (a) Virus quantity by days post inoculation and (b) peak quantity before and during respiratory signs in animals inoculated intratracheally with NiV_M_ and NiV_B_^26, 27^ (N=9). (c) Virus quantity by days post inoculation and (d) peak quantity before and during respiratory signs in animals inoculated intranasally/intratracheally^28, 29^ with NiV_M_ and HeV (N=11). Quantities are shown as log genome equivalents. Error bars represent mean and standard error. Four African green monkeys were excluded from b and d as no samples were positive by PCR and/or the animals did not develop respiratory signs.

#### Ferret studies

Four ferret studies, including 21 animals ^30–33^, were included in the analysis. We were unable to compare virus quantities across studies given a lack of comparable individual-level measurements of virus quantity in the published literature. However, 2 studies directly compared virus quantities at the group-level. Leon and colleagues^31^ inoculated groups of 4 ferrets intranasally with 5,000 50% tissue culture infectious dose (TCID_50_, approximately 3,500 PFU) of NiV_B_, NiV_M_, and HeV. On day 5 post inoculation, animals inoculated with NiV_B_ had the lowest mean virus quantity in nasal samples and animals inoculated with HeV had the highest mean virus quantity in oral samples. Animals inoculated with NiV_M_ had the lowest virus quantity in oral samples on days 6 and 7 post inoculation^31^. Clayton et al^30^ oronasally inoculated 8 ferrets with NiV_B_ and 7 ferrets with NiV_M_, all with 5,000 TCID_50_. They found no statistically significant difference in virus quantity over time in nasal samples (1 to 8 days post inoculation) but observed approximately 10-fold higher virus quantity in oral samples on days 5-6 and days 7-8 post inoculation among ferrets inoculated with NiV_B_. All but one animal in this study had culturable virus in their oral samples on the day of euthanasia.

Although we were unable to make individual-level comparisons of virus quantity, 3 of the ferret studies provided individual-level data on viral shedding duration across virus strains^30, 32, 33^. In all, 21 animals were inoculated with 5,000 TCID_50_ with 3 different henipavirus strains: 8 with NiV_B_, 11 with NiV_M_, and 2 with HeV. Inconsistent timing and frequency of sampling over the course of infection led to difficulties in comparing time to viral shedding onset and duration of viral shedding between virus strains (Figure 6). However, once an animal began oral shedding, all additional samples had evidence of shedding until the day of euthanasia for all animals studied. The time point for euthanasia was based on severity of clinical signs. There were no notable differences in time from inoculation to euthanasia across strains: NiV_B_ (mean, range: 7.6, 7-9), NIV_M_ (8, 5-10), and HeV (7, 7-7). Daily individual-level clinical signs were not reported in any of four studies. Two animals infected with HeV had oral samples investigated for culturable virus on the day of euthanasia; 1 animal had cultural virus in their sample.^32^

**Figure 6.**
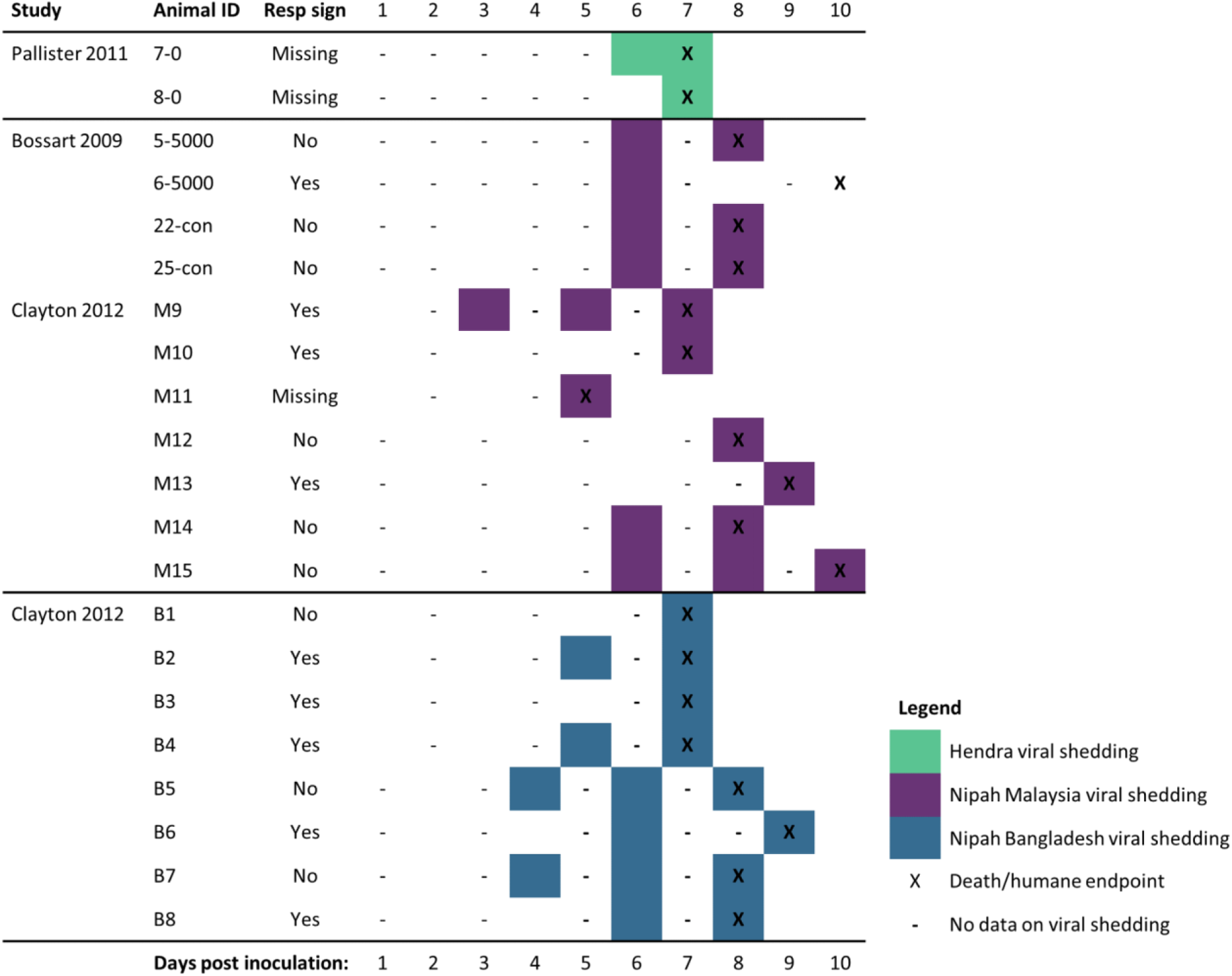
Comparing duration of oral viral shedding, respiratory signs, and timing of humane endpoints among ferrets (N=21) infected with Hendra virus, Nipah virus Bangladesh (NiV_B_), or Nipah virus Malaysia (NiV_M_), by study.

## Discussion

We know very little about the diversity of zoonotic henipaviruses in nature. A new henipavirus infecting shrews and humans, but with no evidence of person-to-person transmission, was reported from China in 2022^34^. In 2021, a new Hendra virus was identified from fruit bats in Australia that had been previously missed by surveillance because the genetic differences in the virus made it undetectable by the PCR diagnostics routinely used^35^. Given the threat these viruses pose for human health, we should aim to understand as best we can the ecology of these viruses and the biological and social determinants of transmission and transmission potential. Our systematic review provides a summary of the evidence thus far, both about what we know and do not know about person-to-person transmission of henipaviruses, as well as the major gaps in how we investigate this phenomenon.

There is substantially more evidence demonstrating the transmission potential of NiV_B_, compared to NiV_M_ or HeV, from both the studies of human epidemiology and animal infection. Our findings suggest that transmission likely occurs through viral shedding from the respiratory tract, with additional evidence of shedding in urine and semen. Two studies have demonstrated environmental contamination with viral RNA^19, 36^, but the role of environmental contamination in transmission remains unknown. However, the role of viral shedding in urine or semen in contributing to onward transmission remains unexplored. The epidemiologic evidence suggests that the heterogeneity of pathogenesis, including the amount of virus being shed and clinical respiratory disease, determines the frequency of super-spreading events. Case-patients infected with NiV_B_ in both India and Bangladesh who transmitted infection experienced more severe symptoms, particularly respiratory distress, all died, and exhibited a shorter time to death from illness onset than case-patients who did not transmit to others. If transmission between people is driven by the amount of virus shed in respiratory secretions and only a few case-patients transmit the virus to the majority of secondary cases, then we would expect high variation in the amount of virus shed by individuals as a major driver of human transmission potential. The substantial variation in the amount of virus shed between African green monkeys infected with NiV_B_ in respiratory secretions supports the conclusion that differences in the number of cases each person infects may be primarily driven by the amount of virus they shed, which could be influenced by biological features of the individual and/or the virus strain.

Our review also suggests that NiV_M_ poses a risk for person-to-person spread. First, there is evidence that NiV_M_ has been transmitted from person to person, both from anecdotal evidence in the case of a family caregiver with relapsing encephalitis^17^ and demonstrated transmission in the Philippines outbreak. Given that less than 1 in 10 patients infect someone else in settings where person-to-person transmission does occur, and that most secondary cases reported have been family caregivers, it is possible that transmission between people simply went unnoticed in Malaysia or Singapore where this type of transmission between the case-patients was not actively investigated. Although non-human primates infected with NiV_M_ did not shed as much virus as those infected with NiV_B_, they were symptomatic and shed virus longer, which could also pose a transmission advantage. The Philippines outbreak was small but showed epidemiologic features of person-to-person transmission very similar to outbreaks in Bangladesh and India, including the proportion of cases who transmitted to others, and the relative frequency of adult males with respiratory symptoms as transmitters. The proportion of case-patients with respiratory symptoms was greater in the Philippines than in Malaysia, possibly suggesting phenotypic diversity among the NiV_M_ strain.

The potential for HeV to be transmitted between people is less clear but should not be discounted. With only 6 human cases of HeV reported in the literature and 7 human cases ever reported, there are too few observations to determine if there are real differences in the transmission potential in humans compared to NiV_B_ or NiV_M_. In addition, most African green monkeys infected with HeV shed virus at similar times, and with accompanying respiratory signs, compared to monkeys infected with NiV_B_ or NiV_M._ Furthermore, animals infected with HeV shed significantly higher titers of virus in respiratory specimens than animals infected with NiV_M_ in the same study. If shedding high amounts of virus in respiratory secretions is important for transmission between people, then HeV may be more transmissible than NiV_M._

Twenty-seven years have passed since the first human infection with a henipavirus was reported^15^, and there are >600 human infections reported in the literature, yet only 40 have had data investigating viral shedding reported. Many studies did not investigate transmission to household contacts^7, 16^, and 1 outbreak in Siliguri, India, where nosocomial transmission was a key driver of the outbreak, provides too few details to recreate transmission chains or identify superspreading events^37^. Given that nosocomial spread has fueled many infectious disease outbreaks, understanding the details of transmission within healthcare settings is critical to designing effective containment and infection control strategies. Moreover, if we want to improve our understanding of the transmission potential of henipaviruses and distinguish the characteristics of these highly fatal viruses that might contribute to differential transmission dynamics, we need a systematic approach to human epidemiologic and clinical investigations and animal models. Standardized protocols to investigate person-to-person spread and biological and behavioral risk factors for transmission, including the potential for sexual transmission, across henipaviruses and settings would leverage opportunities to learn about these emerging viruses. Though a recent summary of person-to-person studies in Bangladesh provides a good start to this work^1^, more details about each case-patient would be useful to increase our ability to understand transmission potential.

Outbreak investigations should report as much granularity as possible on the route of exposure, timeline of infection and symptoms (e.g. incubation period), paired with serial oral/respiratory and blood specimens with viral load (or Ct values as an indicator), evaluation of other potentially infectious body fluids (e.g. tears, semen, vaginal fluids), case fatality ratio (CFR), timing of death, proportion of transmitters, geographic distribution, spillover frequency, and cluster size^38^. The potential for transmission can be quantified by then establishing such measures as the secondary attack rate, duration of viral shedding, quantity of virus shed, and clinical signs and symptoms that promote transmission of the virus to others. Therefore, publishing detailed line lists from outbreaks could help to build a global database of henipavirus transmission that could be leveraged to improve our collective understanding over time.

The primary reason that henipaviruses are a concern for public health is the possibility that a more highly transmissible strain could emerge. Experimental infections of animals can provide unique opportunities to compare henipaviruses in terms of pathology and viral shedding in a standardized way that can help contextualize the highly imperfect observational epidemiologic data about transmission to assess public health risk. For example, the shedding of virus by African green monkeys prior to onset of overt signs of disease may suggest that transmission by humans could occur prior to symptom onset, though these patterns may also be artifacts of the extremely high infectious doses administered in animal infection models. However, the published experimental studies we identified in our review were largely unsuitable for providing meaningful comparisons because of differences in inoculation routes and titers, and frequency of and methods for measuring viremia and viral shedding. Moreover, these studies, as with the human studies, relied primarily on measurements of viral load using PCR, which does not differentiate viable from non-viable virus. These studies, including assessments of virus viability, are costly and time consuming and we should maximize their ability to inform our understanding of transmission through greater harmonization of protocols across labs and animal models. Increasing the diversity of virus strains used in these studies would provide additional insights into the role of viral genetics in transmission potential.

Though our review highlights NiV_B_ as a greater threat to humans compared to the other henipavirus strains, we cannot dismiss the risk that the other known and yet undiscovered viruses may pose. As the family of known henipaviruses continues to grow, shared protocols for human investigations and animal experiments are urgently needed to capitalize on more opportunities to advance our understanding of transmission risk.

## Author contributions

ESG conceived of the study idea; ESG and KHL worked with an informationist to design the search strategy; KHL, STH, AS, and FKJ performed the analyses; all authors reviewed studies for inclusion in the analysis, extracted data from studies, and contributed to manuscript writing.

## Materials and communications

Correspondence related to this paper should be addressed to Emily S. Gurley.

## Data Availability

All data used in this study are publicly available and the extracted data are available from study authors are available upon reasonable request.

---

**Supplementary Figure 1.**
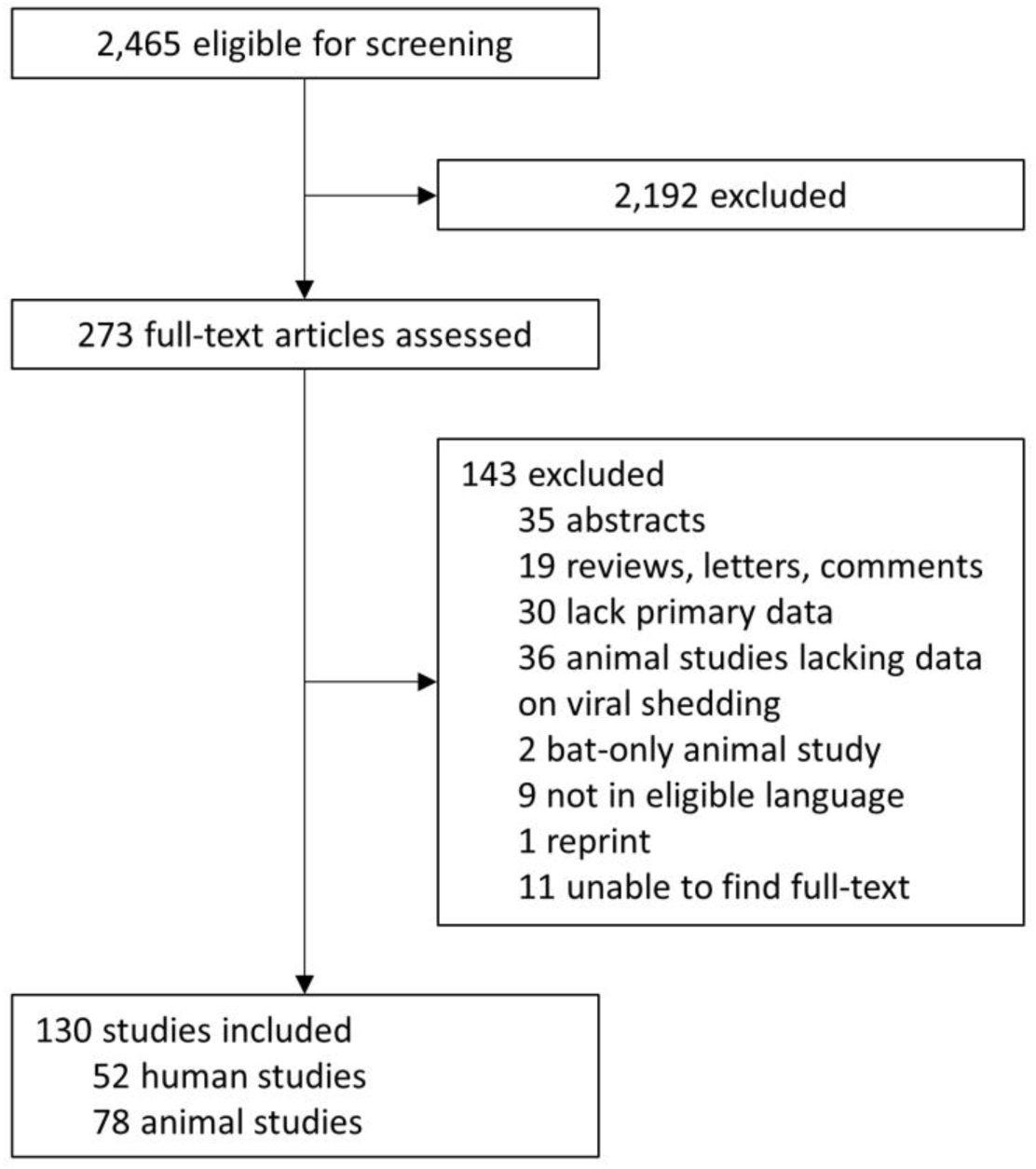
**Study selection**

**Supplementary Figure 2.**
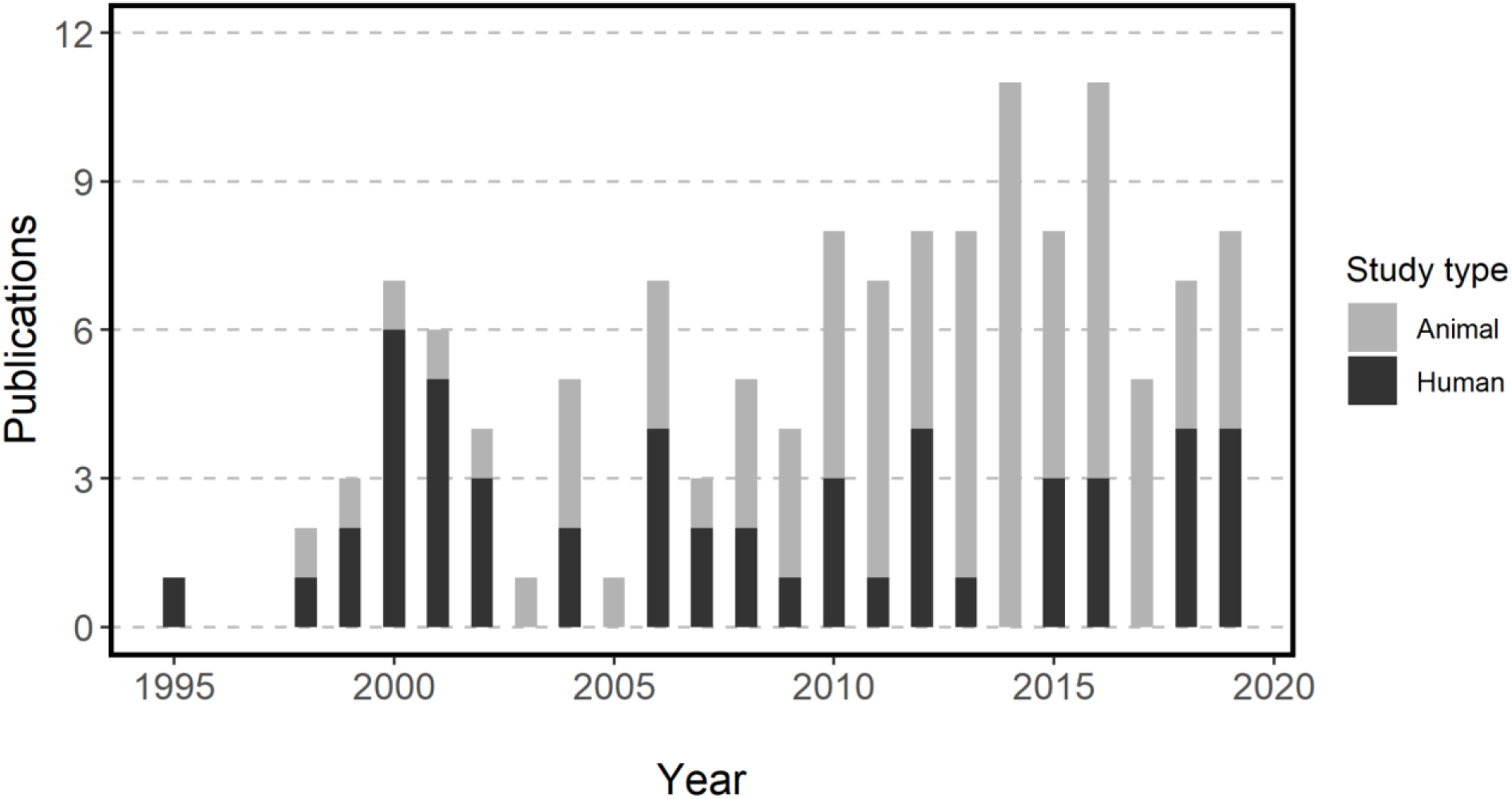
Studies included in systematic review, by study type and year of publication.

**Supplementary Figure 3.**
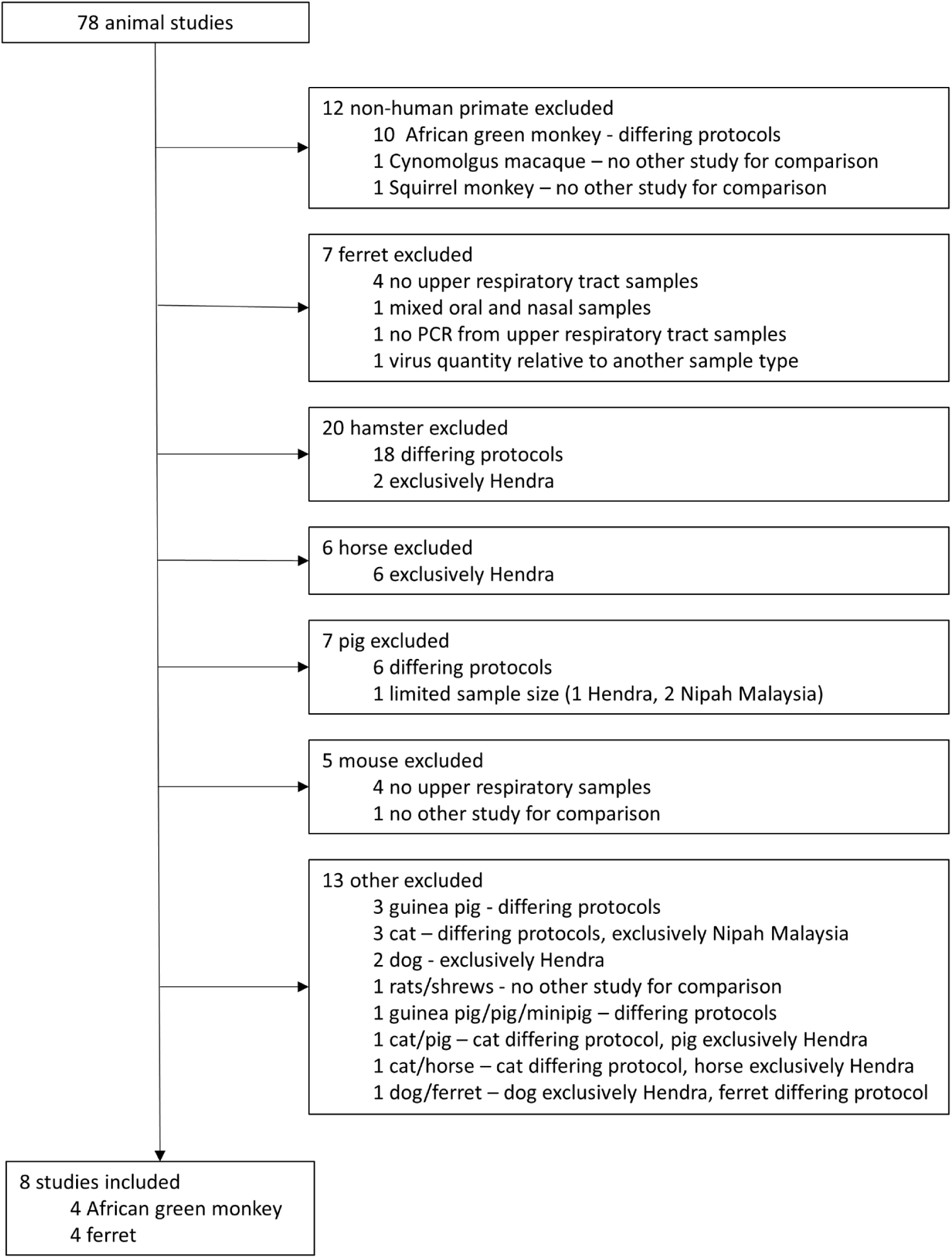
Reasons for animal study exclusion.

**Supplementary Figure 4.**
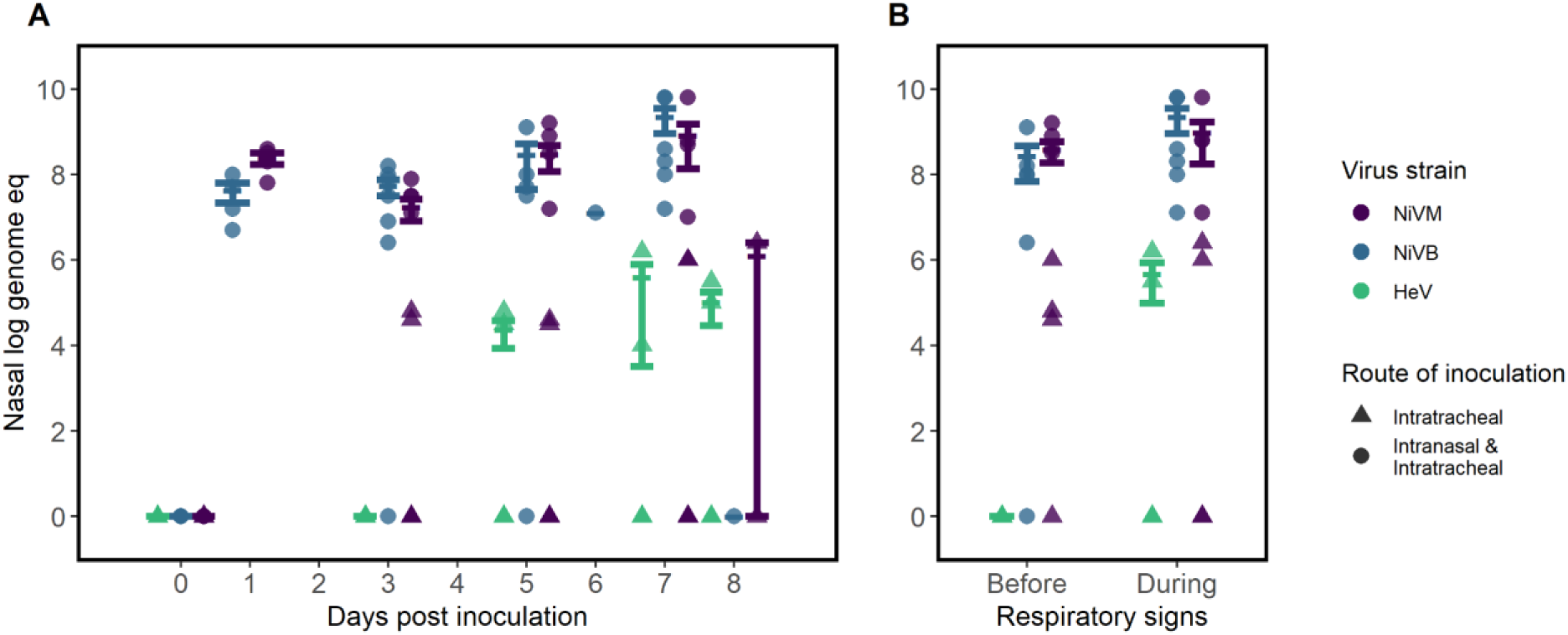
Virus quantity in nasal samples of African green monkeys inoculated in studies with similar study protocols. Quantities are shown as log genome equivalents by day post inoculation (A) and as the peak value before and during respiratory signs (B). Error bars represent mean and standard error. Four African green monkeys were excluded from B as no samples were positive by PCR and/or did not develop respiratory signs.

**Supplementary Table 1.**
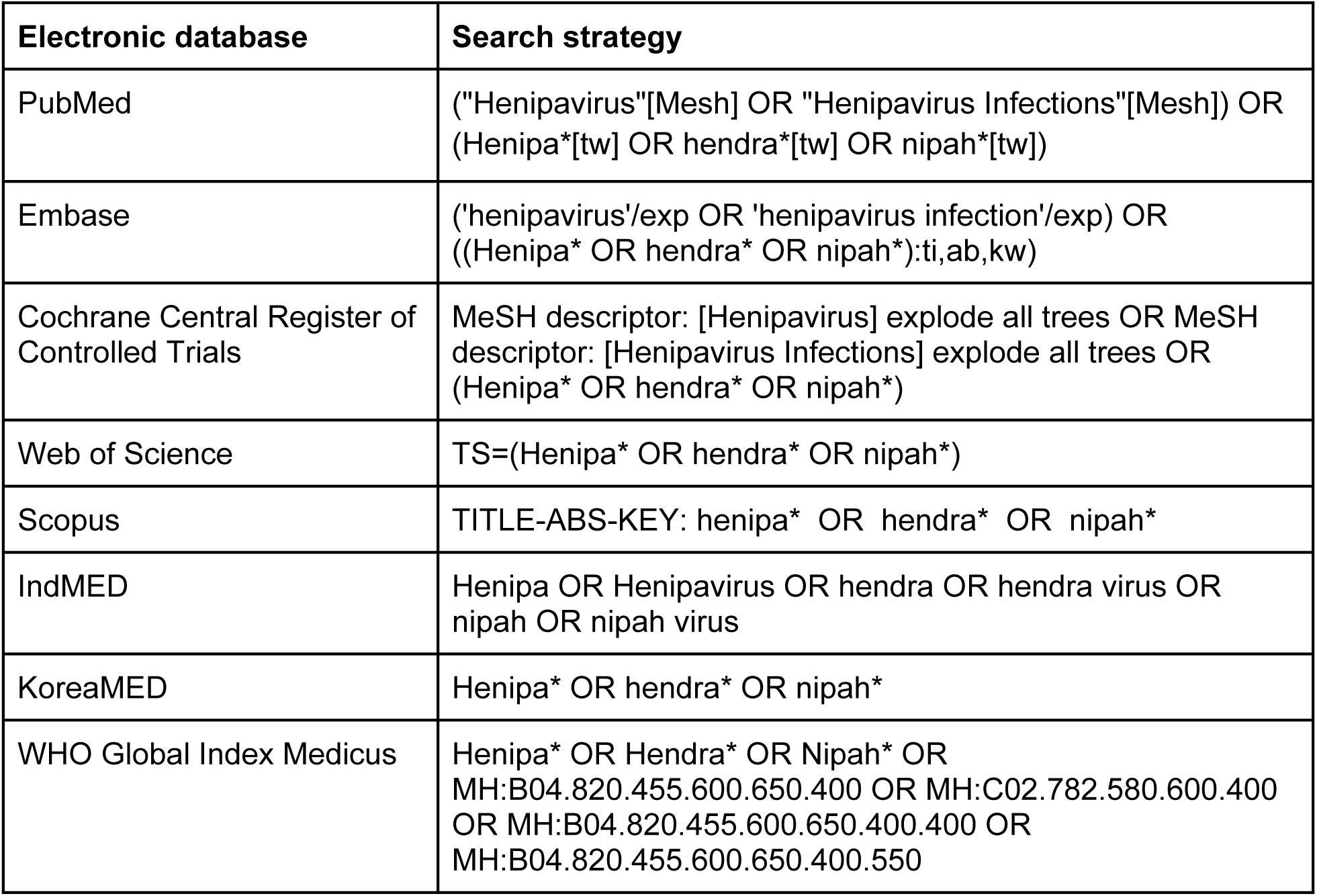
Electronic databases and search strategies

**Supplementary Table 2.**
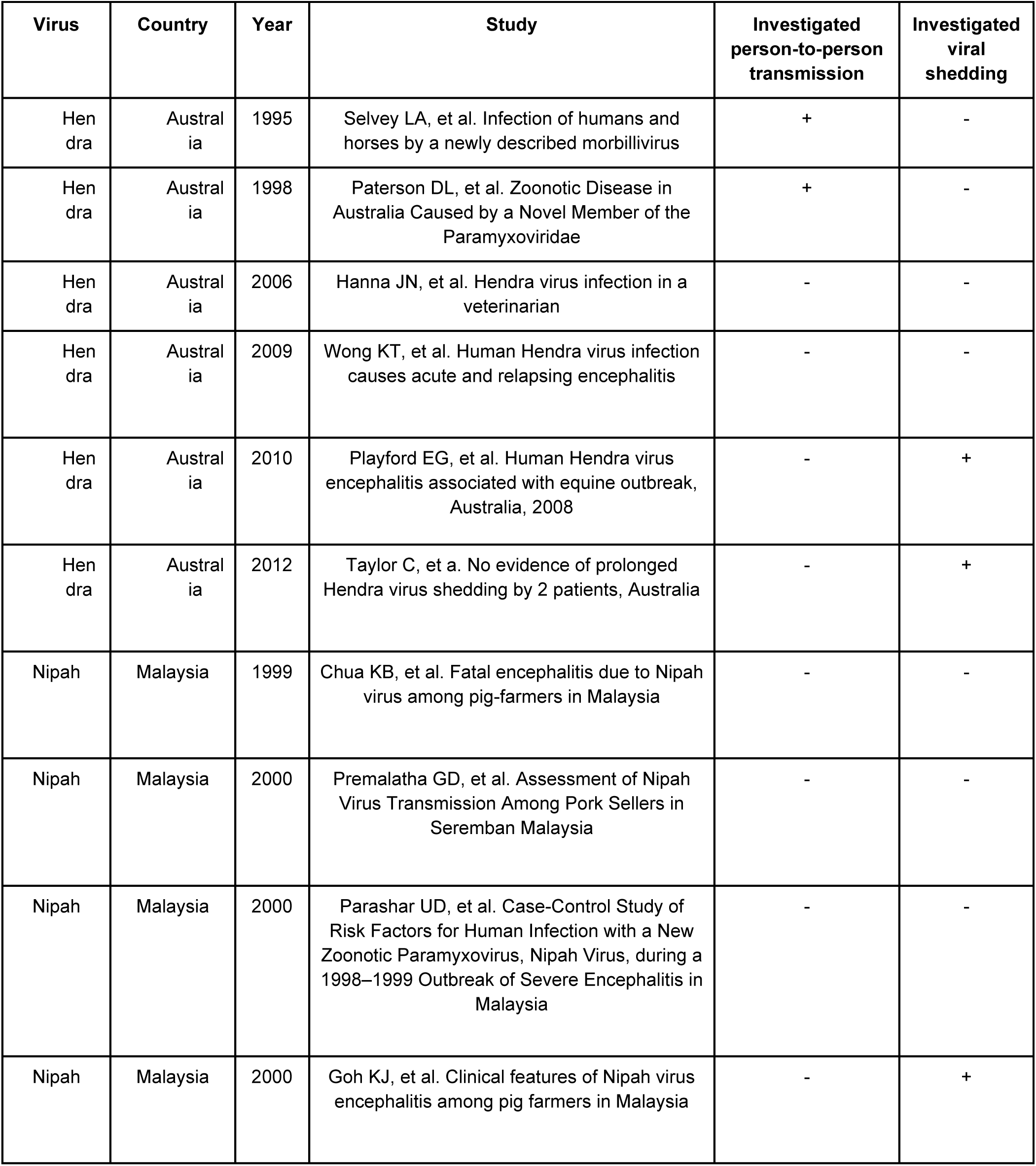
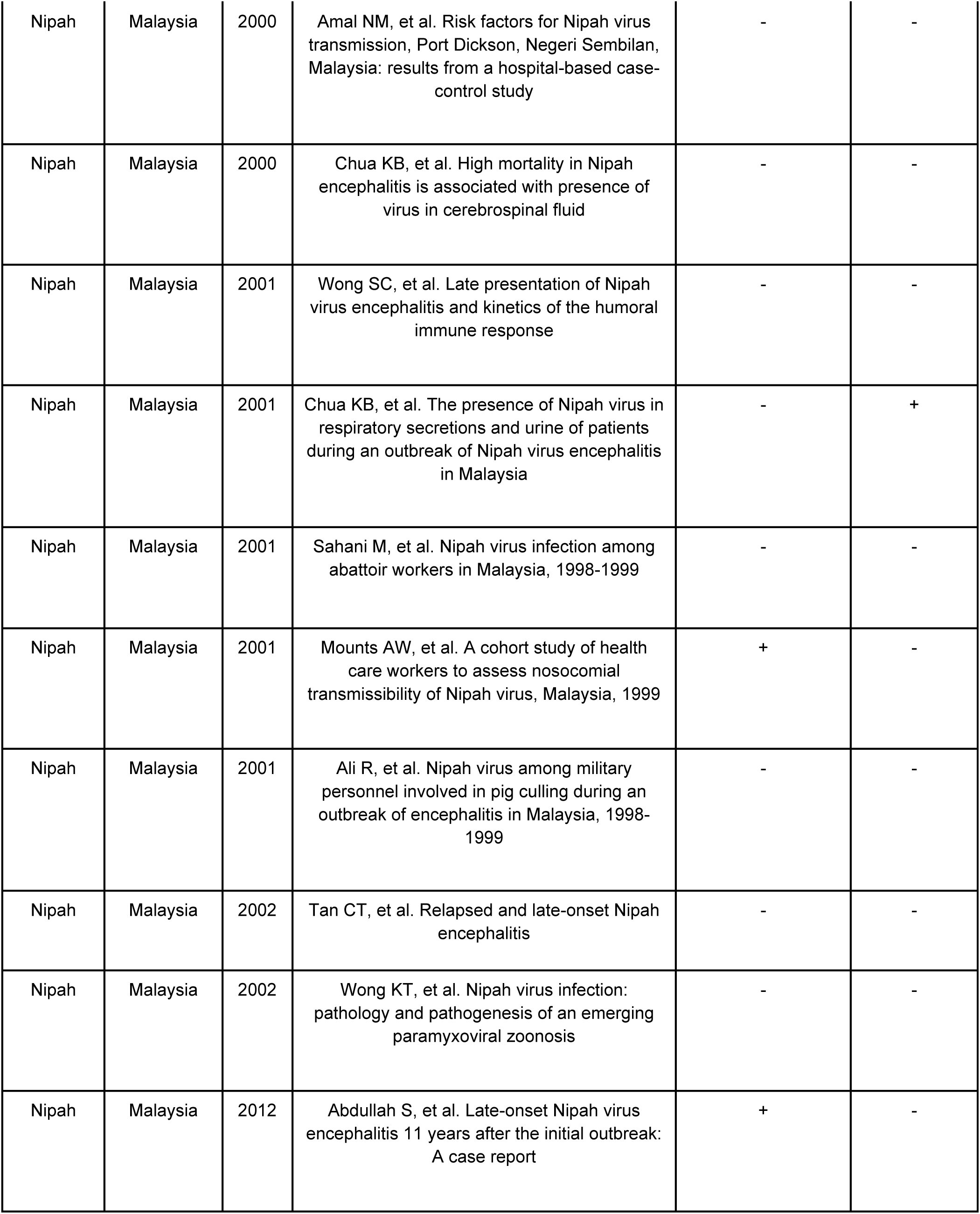
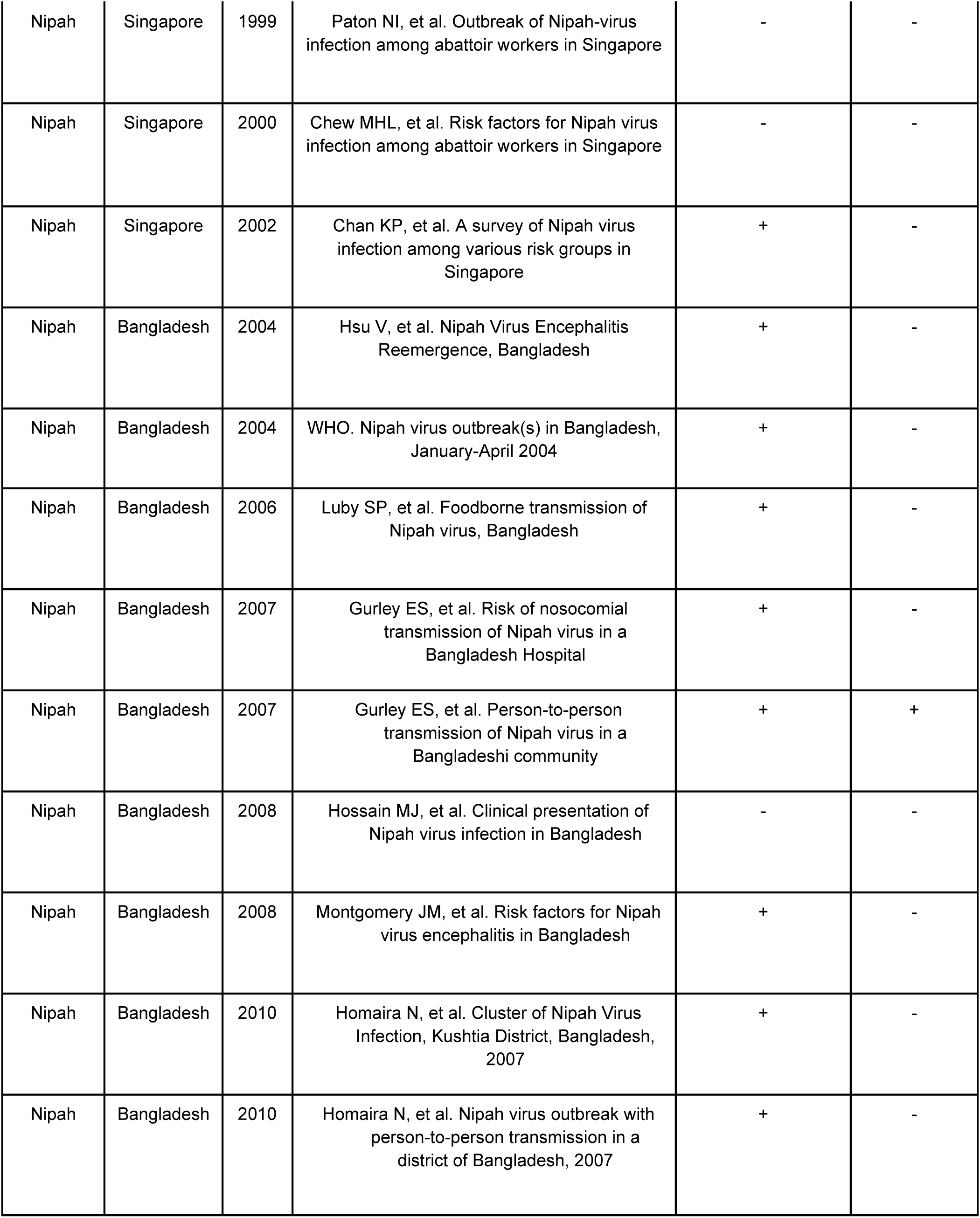
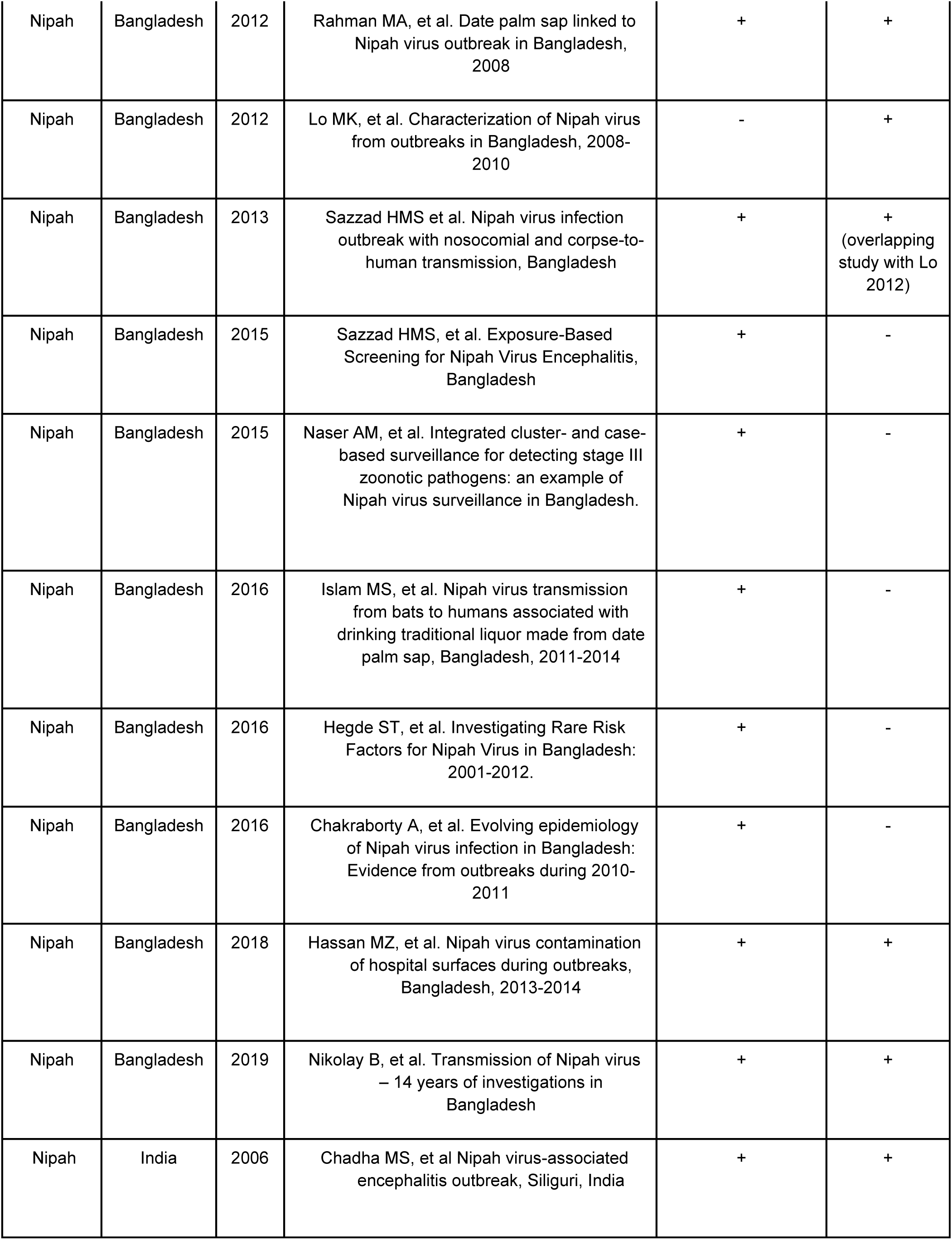
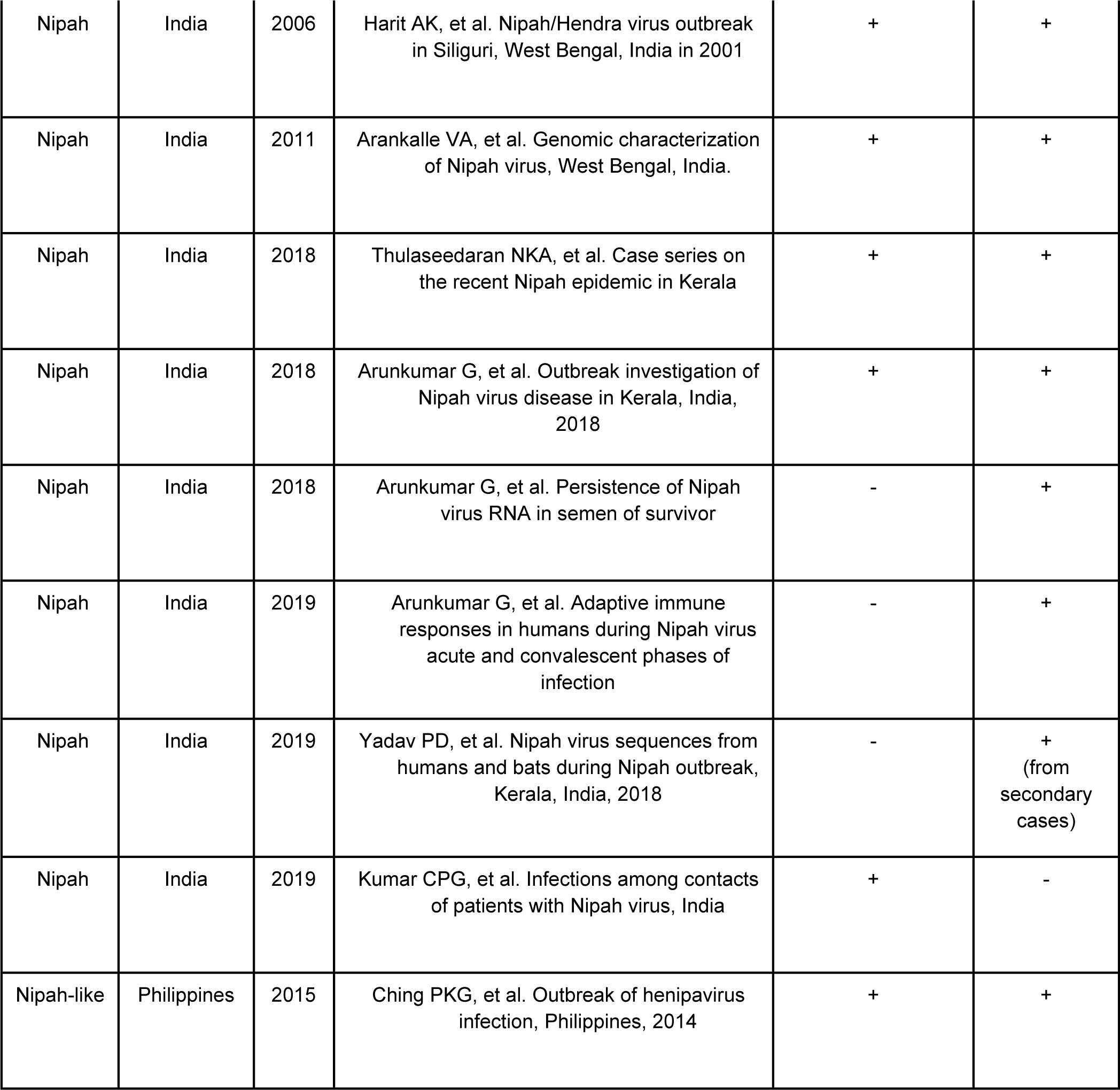
Studies published through May 2019 on human henipavirus infections, by year and country (N=52)

**Supplementary Table 3.**
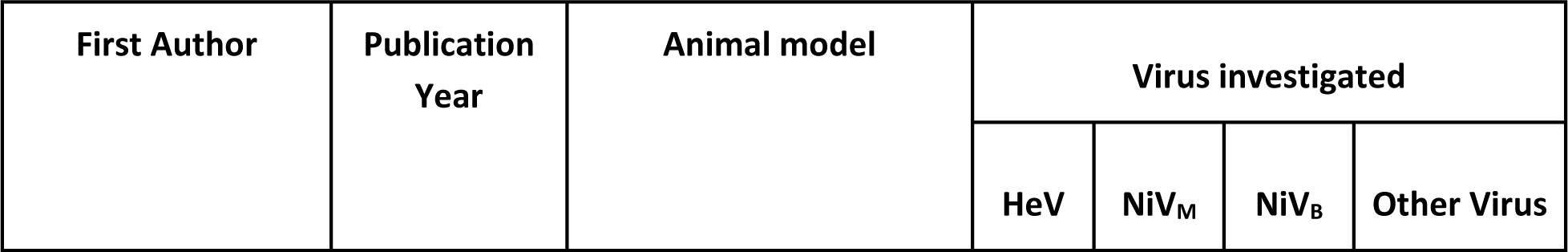
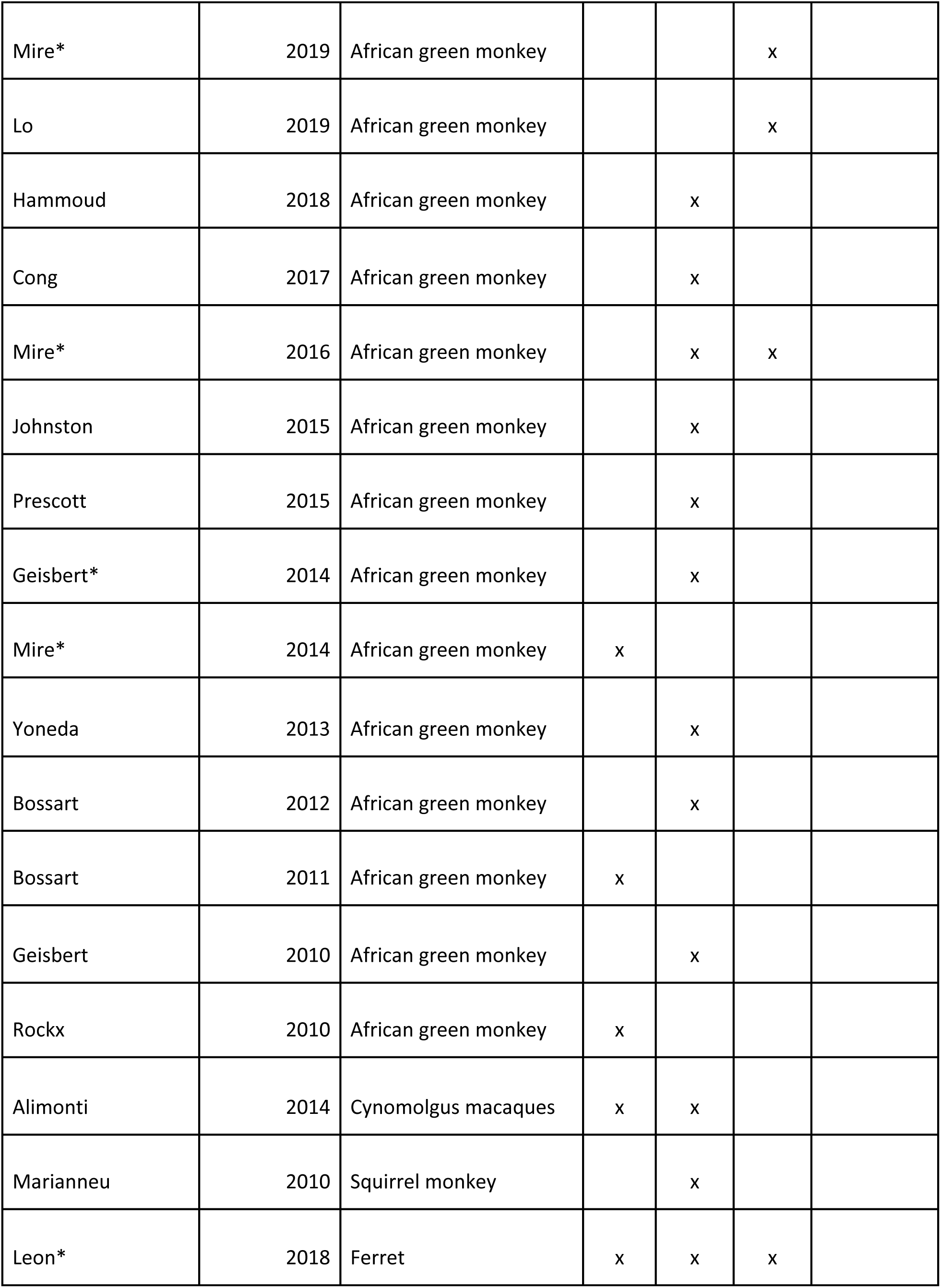
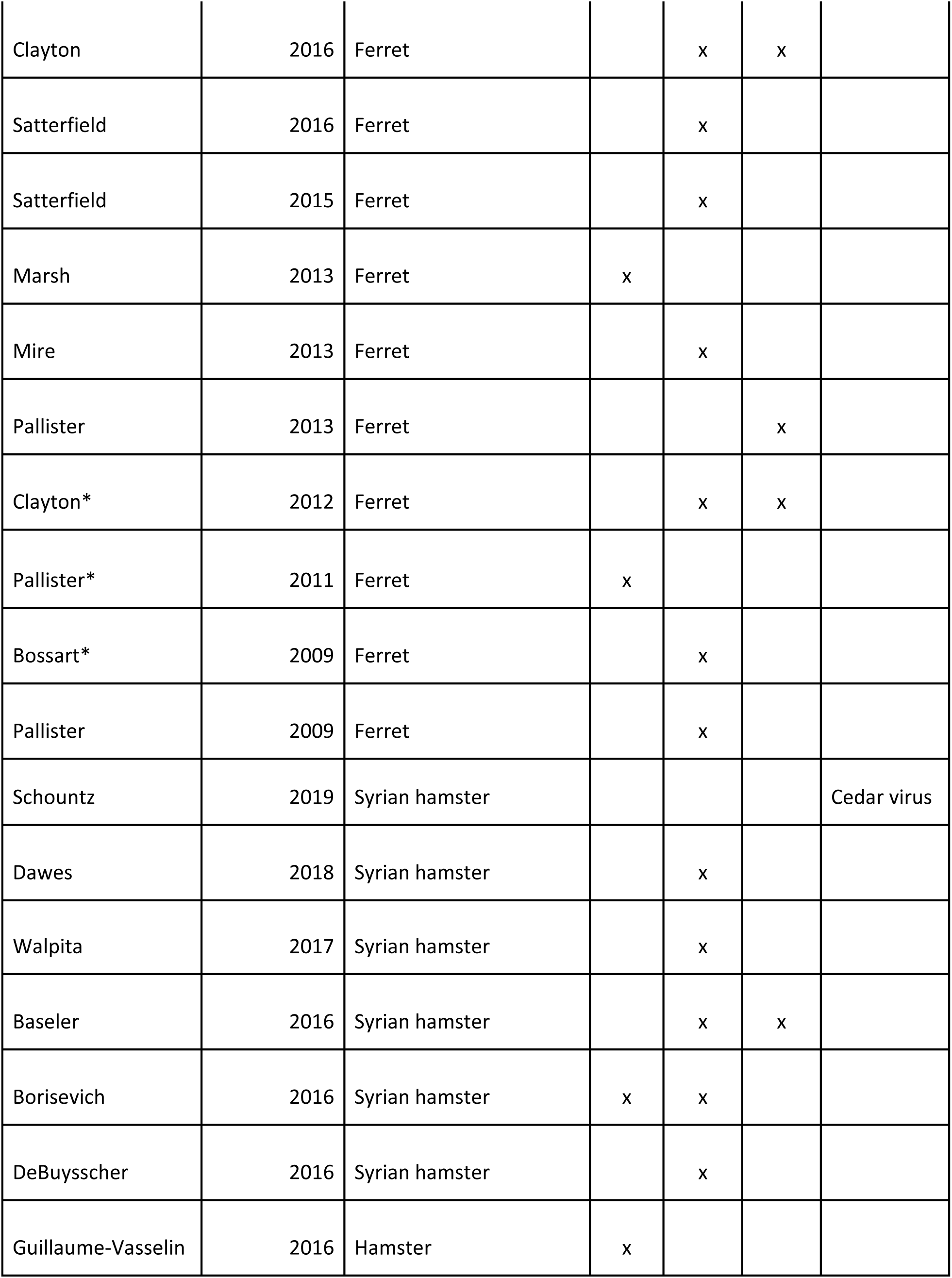
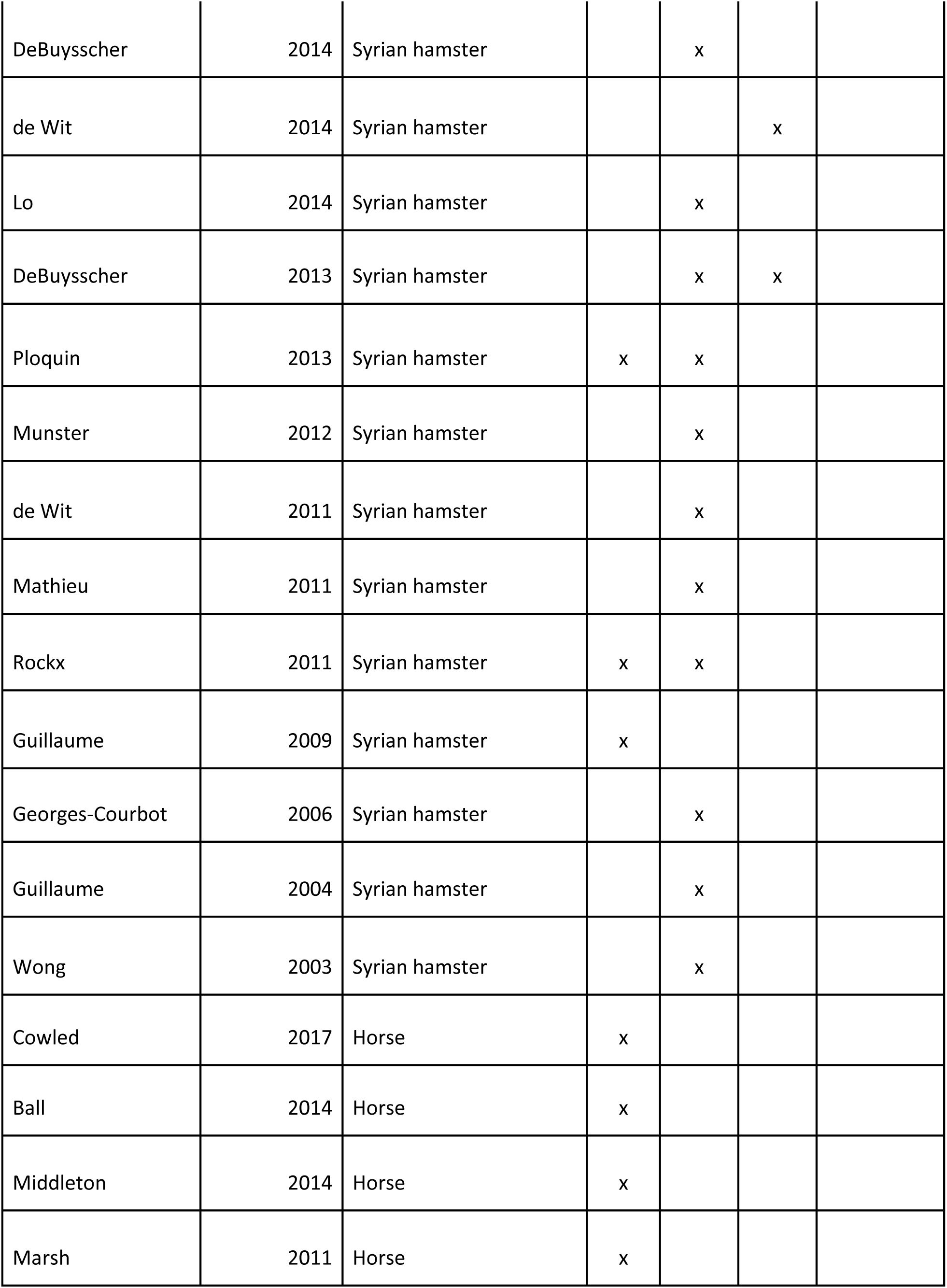
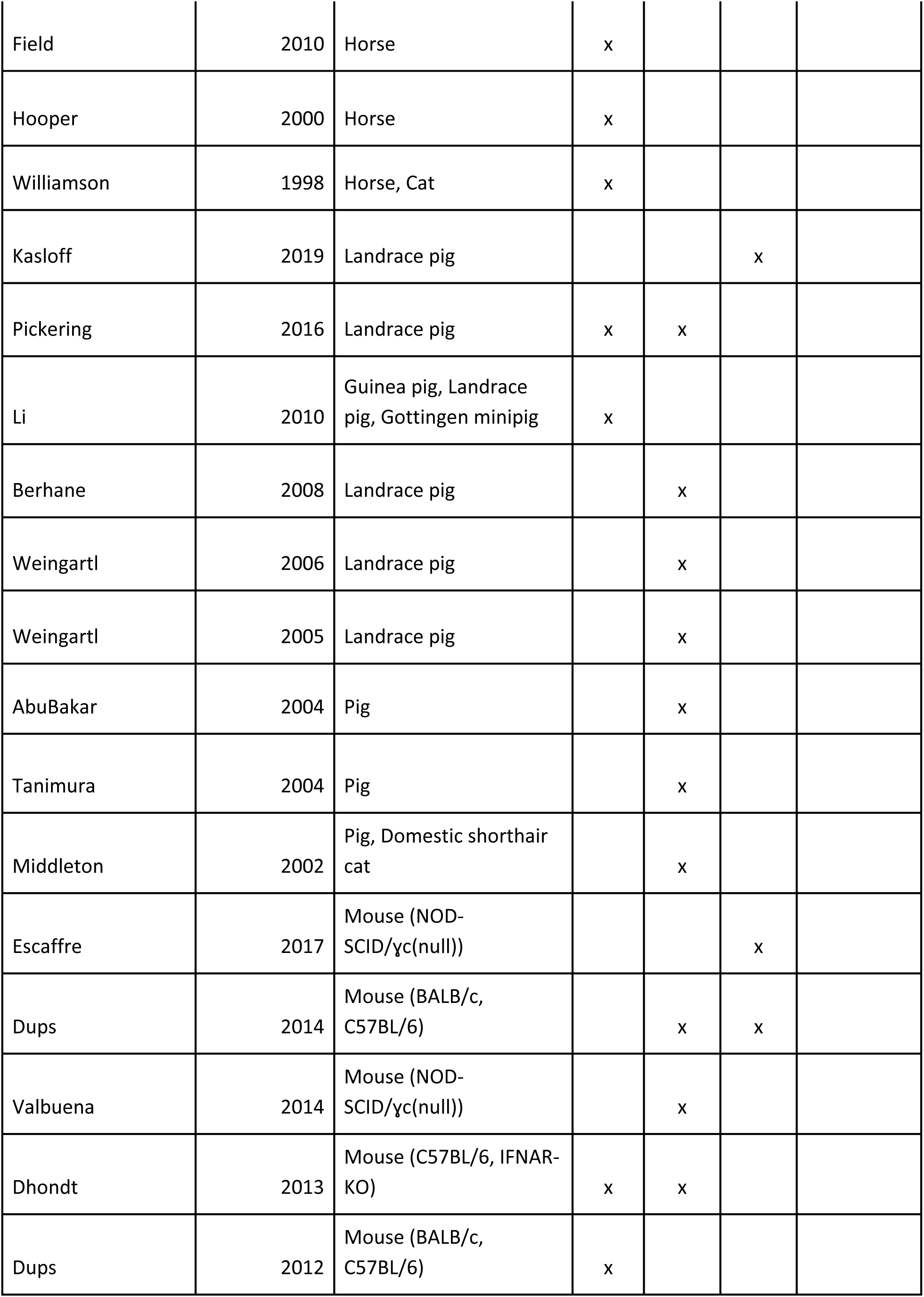
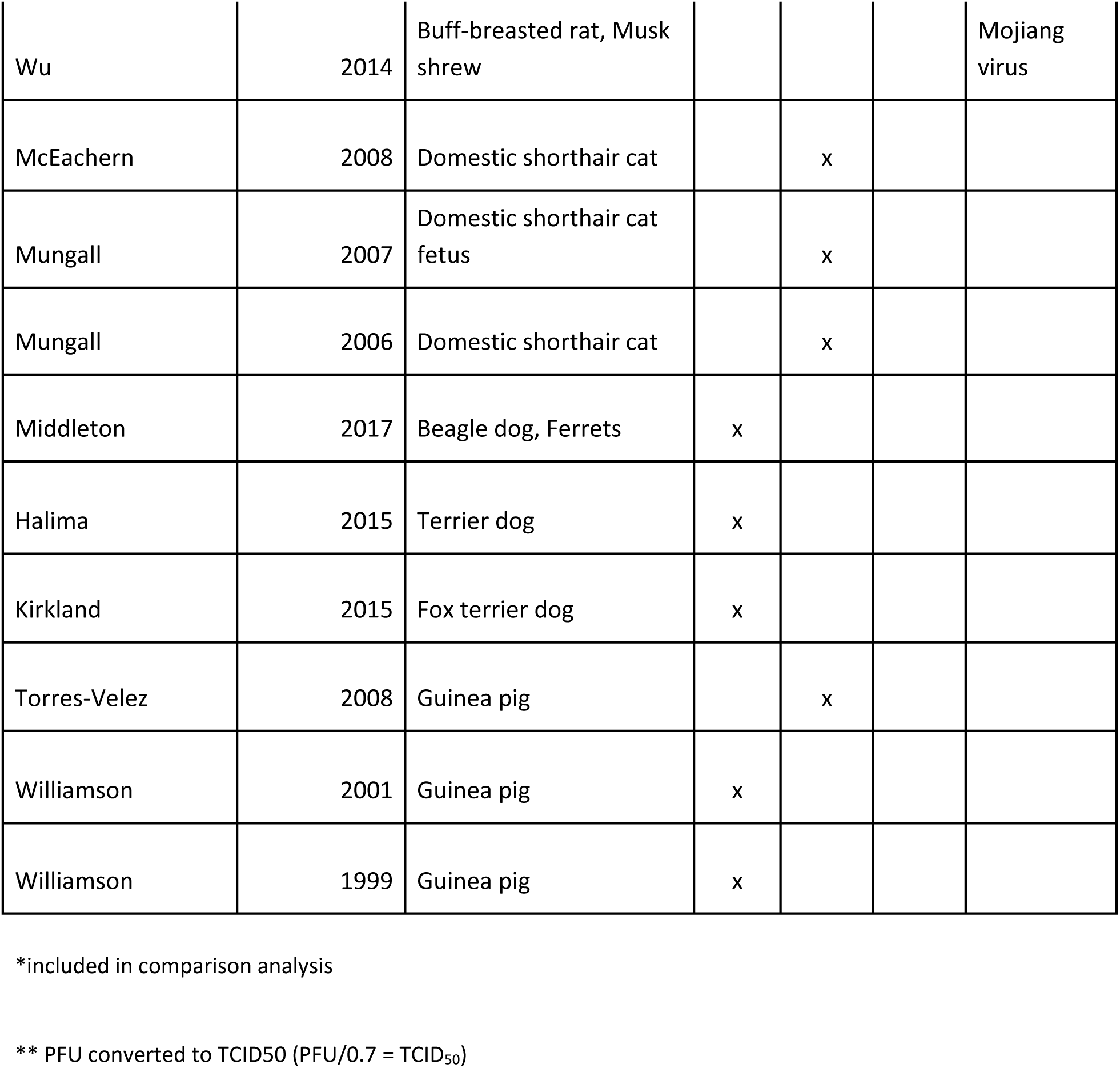
Studies published through May 2019 on animal henipavirus infections, by first author name, year of publication, and animal model (N=78)

## References

1. Nikolay, B. et al. Transmission of Nipah Virus—14 Years of Investigations in Bangladesh. N. Engl. J. Med. 380, 1804–1814 (2019).

2. Luby, S. P. & Gurley, E. S. Epidemiology of henipavirus disease in humans. Curr. Top. Microbiol. Immunol. 359, 25–40 (2012).

3. Marsh, G. A. et al. Cedar virus: a novel Henipavirus isolated from Australian bats. PLoS Pathog. 8, e1002836 (2012).

4. Mounts, A. W. et al. A cohort study of health care workers to assess nosocomial transmissibility of Nipah virus, Malaysia, 1999. J. Infect. Dis. 183, 810–813 (2001).

5. Whitmer, S. L. M. et al. Inference of Nipah virus evolution, 1999-2015. Virus Evol 7, veaa062 (2021).

6. Harcourt, B. H. et al. Genetic characterization of Nipah virus, Bangladesh, 2004. Emerg. Infect. Dis. 11, 1594–1597 (2005).

7. Ching, P. K. G. et al. Outbreak of henipavirus infection, Philippines, 2014. Emerg. Infect. Dis. 21, 328–331 (2015).

8. Lau, L. L. H. et al. Viral shedding and clinical illness in naturally acquired influenza virus infections. J. Infect. Dis. 201, 1509–1516 (2010).

9. Quinn, T. C. et al. Viral load and heterosexual transmission of human immunodeficiency virus type 1. Rakai Project Study Group. N. Engl. J. Med. 342, 921–929 (2000).

10. Hanna, J. N. et al. Hendra virus infection in a veterinarian. Medical Journal of Australia vol. 185 562–564 Preprint at https://doi.org/10.5694/j.1326-5377.2006.tb00692.x (2006).

11. Paterson, D. L., Murray, P. K. & McCormack, J. G. Zoonotic disease in Australia caused by a novel member of the paramyxoviridae. Clin. Infect. Dis. 27, 112–118 (1998).

12. Wong, K. T. et al. Human Hendra virus infection causes acute and relapsing encephalitis. Neuropathol. Appl. Neurobiol. 35, 296–305 (2009).

13. Summary of human cases of Hendra virus infection - Control guidelines. https://www.health.nsw.gov.au/Infectious/controlguideline/Pages/hendra-case-summary.aspx.

14. Playford, E. G. et al. Human Hendra virus encephalitis associated with equine outbreak, Australia, 2008. Emerg. Infect. Dis. 16, 219–223 (2010).

15. Selvey, L. A. et al. Infection of humans and horses by a newly described morbillivirus. Med. J. Aust. 162, 642–645 (1995).

16. Chan, K. P. et al. A survey of Nipah virus infection among various risk groups in Singapore. Epidemiol. Infect. 128, 93–98 (2002).

17. Abdullah, S., Chang, L.-Y., Rahmat, K., Goh, K. J. & Tan, C. T. Late-onset Nipah virus encephalitis 11 years after the initial outbreak: A case report. Neurology Asia 17, (2012).

18. Arunkumar, G. et al. Outbreak Investigation of Nipah Virus Disease in Kerala, India, 2018. J. Infect. Dis. 219, 1867–1878 (2019).

19. Gurley, E. S. et al. Person-to-person transmission of Nipah virus in a Bangladeshi community. Emerg. Infect. Dis. 13, 1031–1037 (2007).

20. Yadav, P. D. et al. Nipah Virus Sequences from Humans and Bats during Nipah Outbreak, Kerala, India, 2018. Emerg. Infect. Dis. 25, 1003–1006 (2019).

21. Lee, K. H. et al. Changing Contact Patterns Over Disease Progression: Nipah Virus as a Case Study. J. Infect. Dis. 222, 438–442 (2020).

22. Schountz, T. et al. Differential Innate Immune Responses Elicited by Nipah Virus and Cedar Virus Correlate with Disparate In Vivo Pathogenesis in Hamsters. Viruses 11, (2019).

23. Wu, Z., et al. Novel Henipa-like virus, Mojiang Paramyxovirus, in rats, China, 2012. Emerg. Infect. Dis. 20, 1064–1066 (2014).

24. Chua, K. B. et al. Fatal encephalitis due to Nipah virus among pig-farmers in Malaysia. Lancet 354, 1257–1259 (1999).

25. Field, H. et al. Hendra virus outbreak with novel clinical features, Australia. Emerg. Infect. Dis. 16, 338–340 (2010).

26. Mire, C. E. et al. A recombinant Hendra virus G glycoprotein subunit vaccine protects nonhuman primates against Hendra virus challenge. J. Virol. 88, 4624–4631 (2014).

27. Geisbert, T. W. et al. Therapeutic treatment of Nipah virus infection in nonhuman primates with a neutralizing human monoclonal antibody. Sci. Transl. Med. 6, 242ra82 (2014).

28. Mire, C. E. et al. Pathogenic Differences between Nipah Virus Bangladesh and Malaysia Strains in Primates: Implications for Antibody Therapy. Sci. Rep. 6, 30916 (2016).

29. Mire, C. E. et al Use of Single-Injection Recombinant Vesicular Stomatitis Virus Vaccine to Protect Nonhuman Primates Against Lethal Nipah Virus Disease. Emerg. Infect. Dis. 25, 1144–1152 (2019).

30. Clayton, B. A. et al. Transmission routes for nipah virus from Malaysia and Bangladesh. Emerg. Infect. Dis. 18, 1983–1993 (2012).

31. Leon, A. J. et al. Host gene expression profiles in ferrets infected with genetically distinct henipavirus strains. PLoS Negl. Trop. Dis. 12, e0006343 (2018).

32. Pallister, J. et al. A recombinant Hendra virus G glycoprotein-based subunit vaccine protects ferrets from lethal Hendra virus challenge. Vaccine 29, 5623–5630 (2011).

33. Bossart, K. N. et al. A neutralizing human monoclonal antibody protects against lethal disease in a new ferret model of acute nipah virus infection. PLoS Pathog. 5, e1000642 (2009).

34. Zhang, X.-A. et al. A Zoonotic Henipavirus in Febrile Patients in China. N. Engl. J. Med. 387, 470–472 (2022).

35. Wang, J. et al. A new Hendra virus genotype found in Australian flying foxes. Virol. J. 18, 197 (2021).

36. Hassan, M. Z. et al. Contamination of hospital surfaces with respiratory pathogens in Bangladesh. PLoS One 14, e0224065 (2019).

37. Harit, A. K. et al. Nipah/Hendra virus outbreak in Siliguri, West Bengal, India in 2001. Indian J. Med. Res. 123, 553–560 (2006).

38. Nikolay, B. et al. A Framework to Monitor Changes in Transmission and Epidemiology of Emerging Pathogens: Lessons From Nipah Virus. J. Infect. Dis. 221, S363–S369 (2020).

39. Arankalle, V. A. et al. Genomic characterization of Nipah virus, West Bengal, India. Emerg. Infect. Dis. 17, 907–909 (2011).

40. Ali, R. et al. Nipah Virus Infection Among Military Personnel Involved in Pig Culling during an Outbreak of Encephalitis in Malaysia, 1998-1999. Emerging Infectious Disease journal 7, 759 (2001).

41. Arunkumar, G. et al. Persistence of Nipah Virus RNA in Semen of Survivor. Clin. Infect. Dis. 69, 377–378 (2019).

42. Kumar, C. P. G. et al. Infections among contacts of patients with Nipah virus, India. Emerg. Infect. Dis. 25, 1007–1010 (2019).

43. Luby, S. P., Gurley, E. S. & Jahangir Hossain, M. Transmission of Human Infection with Nipah Virus. Clinical Infectious Diseases vol. 49 1743–1748 Preprint at https://doi.org/10.1086/647951 (2009).

44. Chew, M. H. et al. Risk factors for Nipah virus infection among abattoir workers in Singapore. J. Infect. Dis. 181, 1760–1763 (2000).

45. Goh, K. J. et al. Clinical features of Nipah virus encephalitis among pig farmers in Malaysia. N. Engl. J. Med. 342, 1229–1235 (2000).

46. Wong, K. T. et al. Nipah virus infection: pathology and pathogenesis of an emerging paramyxoviral zoonosis. Am. J. Pathol. 161, 2153–2167 (2002).

